# Cognitive deficits in people who have recovered from COVID-19 relative to controls: An N=84,285 online study

**DOI:** 10.1101/2020.10.20.20215863

**Authors:** Adam Hampshire, William Trender, Samuel R Chamberlain, Amy Jolly, Jon E. Grant, Fiona Patrick, Ndaba Mazibuko, Steve Williams, Joseph M Barnby, Peter Hellyer, Mitul A Mehta

## Abstract

Case studies have revealed neurological problems in severely affected COVID-19 patients. However, there is little information regarding the nature and broader prevalence of cognitive problems post-infection or across the full spread of severity. We analysed cognitive test data from 84,285 Great British Intelligence Test participants who completed a questionnaire regarding suspected and biologically confirmed COVID-19 infection. People who had recovered, including those no longer reporting symptoms, exhibited significant cognitive deficits when controlling for age, gender, education level, income, racial-ethnic group and pre-existing medical disorders. They were of substantial effect size for people who had been hospitalised, but also for mild but biologically confirmed cases who reported no breathing difficulty. Finer grained analyses of performance support the hypothesis that COVID-19 has a multi-system impact on human cognition.

**Significance statement:** There is evidence that COVID-19 may cause long term health changes past acute symptoms, termed ‘long COVID’. Our analyses of detailed cognitive assessment and questionnaire data from tens thousands of datasets, collected in collaboration with BBC2 Horizon, align with the view that there are chronic cognitive consequences of having COVID-19. Individuals who recovered from suspected or confirmed COVID-19 perform worse on cognitive tests in multiple domains than would be expected given their detailed age and demographic profiles. This deficit scales with symptom severity and is evident amongst those without hospital treatment. These results should act as a clarion call for more detailed research investigating the basis of cognitive deficits in people who have survived SARS-COV-2 infection.

## Introduction

There is growing evidence that individuals with severe COVID-19 disease can develop a range of neurological complications^1-3^ including those arising from stroke^4,5^, encephalopathies^6^, inflammatory syndrome^4,7^, microbleeds^4^ and autoimmune responses^8^. There are concerns regarding potential neurological consequences due to sepsis, hypoxia and immune hyperstimulation^4,9,10^, with reports of elevated cerebrospinal fluid autoantibodies in patients with neurological symptoms^11^, white matter change in the brain^2,12,13^, and psychological and psychiatric consequences at the point of discharge^14^. However, it is yet to be established whether COVID-19 infection is associated with cognitive impairment at the population level; and if so, how this differs with respiratory symptom severity and relatedly, hospitalisation status^4,15^. Measuring such associations is challenging. Longitudinal collection of cognitive data from pre-to post-COVID is extremely problematic because infection is unpredictable. Furthermore, it is important to include key minority sub-populations, for example, older adults, racial-ethnic groups, and people with preexisting medical conditions^16-18^. This motivated us to take a large-scale cross-sectional approach, whereby individuals who have recovered from COVID-19 infection were compared to concurrently obtained controls while accounting for the uneven sociodemographic distribution of virus prevalence and the associated population variability in cognition. At the time of writing, we had collected comprehensive cognitive test and questionnaire data from a very large cross-section of the general public, predominantly within the UK, as part of the Great British Intelligence Test - a collaborative project with BBC2 Horizon. Due to the high visibility of the study, this cohort spanned a broad age and demographic range. During May, at the peak of the UK lockdown, we expanded the questionnaire (table S1) to include questions pertaining to the impact of the pandemic, including suspected or confirmed COVID-19 illness, alongside details of symptom persistence and severity, and relevant pre-existing medical conditions. We analysed data from 84,285 individuals (figure 1 & table S2) who completed the full extended questionnaire to determine whether those who had recovered from COVID-19 showed objective cognitive deficits when performing tests of semantic problem solving, spatial working memory, selective attention and emotional processing; and whether the extent and/or nature of deficit related to severity of respiratory symptoms as gauged by level of medical assistance.

**Figure 1.**
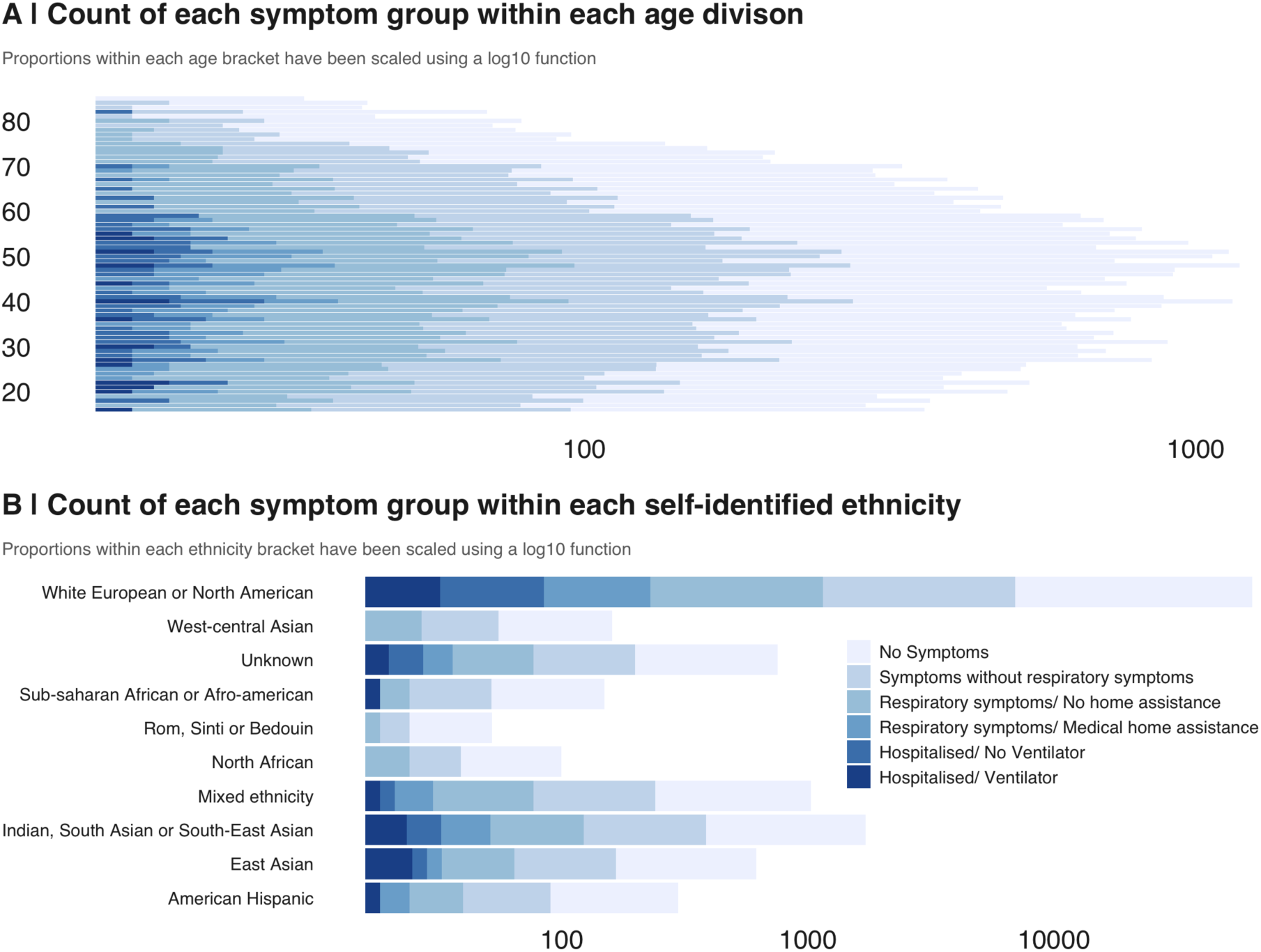
COVID-19 illness in relation to cohort demographics. **A |** Distributions of people reporting having recovered from COVID-19 broken down according to the treatment that they received for respiratory symptoms. Note, the broad and matched age distribution for all sub-groups. **B |** People from a broad range of self-identified ethnic groups took part in this study.

## Results

### Participants

Amongst 84,285 participants, 60 reported being put on a ventilator, a further 147 were hospitalised without a ventilator, 176 required medical assistance at home for respiratory difficulties, 3466 had respiratory difficulties and received no medical assistance and 9201 reported being ill without respiratory symptoms. Amongst these 361 reported having had a positive biological test, including the majority of hospitalised cases. Full details of cohort age and sociodemographic distributions are provided in supplementary tables 2a-i.

### Global cognitive deficits

Generalised linear modelling (GLM) was applied to determine whether global cognitive scores covaried with respiratory COVID-19 symptom severity after factoring out age, sex, handedness, first language, education level, country of residence, occupational status and earnings. There was a significant main effect (F(5,84279)=11.848 p=1.76E-11), with increasing degrees of cognitive underperformance relative to controls dependent on level of medical assistance received for COVID-19 respiratory symptoms (Figure 2a - table S3). People who had been hospitalised showed large-medium scaled global performance deficits dependent on whether they were (−0.57 standard deviations (SDs) N=60) vs. were not (−0.45SDs N=147) put onto a ventilator. Those who remained at home (i.e., without inpatient support) showed small statistically significant global performance deficits (assisted at home for respiratory difficulty −0.12 SD N=176; no medical assistance but respiratory difficulty −0.10 SDs N=3466; ill without respiratory difficulty −0.04 SDs N=9201).

**Figure 2.**
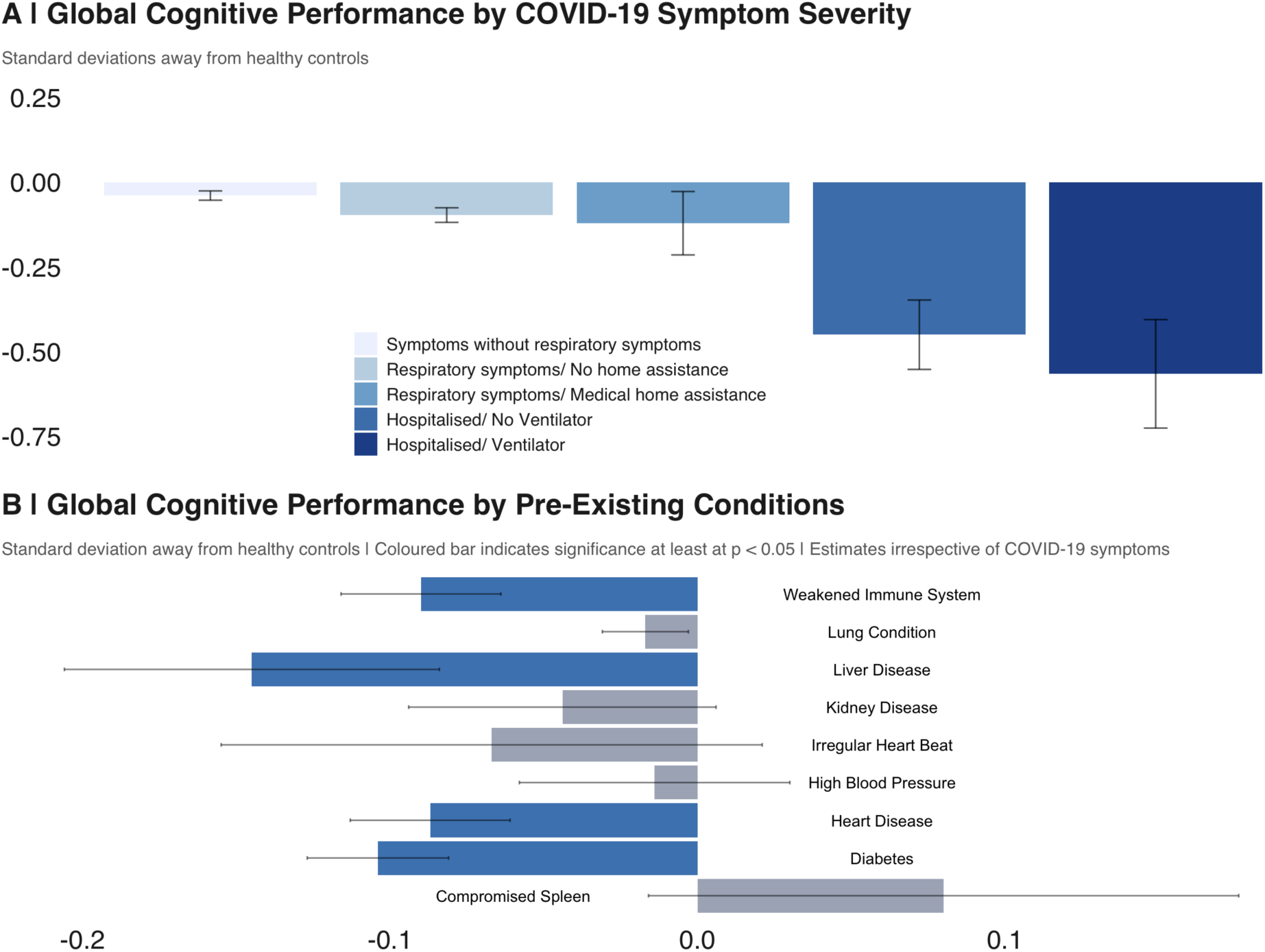
Cognitive deficits in people with suspected and confirmed Covid-19 illness. **A |** People who reported having recovered from COVID-19 performed worse in terms of global score. The scale of this deficit increased with the level of treatment received for respiratory difficulty. **B |** The scale of the deficit associated with COVID-19 was substantially greater than common pre-existing conditions that are associated with vulnerability to the virus and cognitive problems.

### Relationship between cognitive deficits and positive biological test

The GLM was re-estimated including confirmation of COVID-19 by biological test as a main effect (table S4). In proportion with the number of UK confirmed cases, 361 people reported a positive biological test, including 87% of the hospitalised with ventilator sub-group. There were significant main effects of positive test (F(1,84274)=21.624 p=3.32E-06 estimate=-0.33SDs) and respiratory severity (F(5,84274)=7.51 p=4.70E-07). Intriguingly though, the interaction was non-significant (F(4,84274)=0.97 p=0.420), indicating a possible deficit for mild cases who were biologically confirmed as positive for COVID-19. A further GLM restricted to those who reported no breathing difficulties (bio-positive=187 vs. suspected=9014) confirmed this, with a robustly greater global performance deficit for positively confirmed cases (t=-3.49 p<0.0001 estimate=-0.32SDs). Repeating the analysing for people who reported staying at home with breathing difficulty but no assistance (bio-positive=84 suspected=3382) showed a similar scaled deficit (t=-2.611 p=0.009 estimate=-0.36SDs). A larger relationship was evident amongst cases who went to hospital but were not put on a ventilator (bio-positive=24 vs suspected=123, t=-2.401 p=0.018 estimate=0.71SDs).

### Ongoing symptoms and pre-existing conditions

One possibility was that the observed cognitive deficits related to ongoing symptoms of COVID-19 infection, e.g., high temperature or respiratory problems. Only a small proportion (0.76%) of participants reported having residual symptoms, although this did include most (78%) of the ventilator group. When report of residual COVID-19 symptoms was included in the GLM (table S5), the main effect of respiratory severity was undiminished (F(5,84278)=9.55 p=4.03E-09). The main effect of residual symptoms was formally non-significant and of small effect size (F(1,84278)=3.82 p=0.051 estimate −0.10 SDs). Another possibility was that the observed cognitive deficits had a basis in pre-existing conditions. When a GLM was estimated including common pre-existing conditions (Fig. 2b - table S6), a number of them showed the expected association with reduced cognitive performance. However, the statistical significance and scale of the respiratory severity main effect remained approximately the same (F(5,84270)=11.07 p=1.262E-10). Furthermore, the effect size for those who had been hospitalised was of substantially greater scale than the other conditions examined.

### Finer grained analysis of cognitive domains

Finally, the cognitive deficits were examined at a finer grain. First, we applied principal component analysis (PCA) to the test summary scores, producing three components. GLMs showed a robust main effect of respiratory symptom severity for component 1 (Figure 3a - table S7), labelled Semantic Problem Solving (F(5,84279)=5.89 p=2E-05), with significantly scaled deficits for the two hospitalised groups (no ventilator −0.28SDs, ventilator - 0.62SDs). Component 2, labelled Visual Attention, also showed a significant main effect (F(5,84279)=7.46 p=2E-07). This reflected more graded deficits in attention scores, including significantly scaled reductions in performance for the three groups who received medical assistance (at home −0.22SDs, no ventilator −0.33SDs, ventilator −0.33SDs) and small but statistically significant deficits for the milder groups (respiratory symptoms −0.07SDs, no respiratory symptoms −0.06SDs). Component 3, labelled Spatial Working Memory, showed a threshold level main effect(F(5,84279)=2.23 p=0.049), with the deficit for the more severe hospital group being statistically non-significant. Analysis of individual test scores (Figure 3b and table S8) further highlighted this broad but variable profile of deficits across cognitive domains.

**Figure 3.**
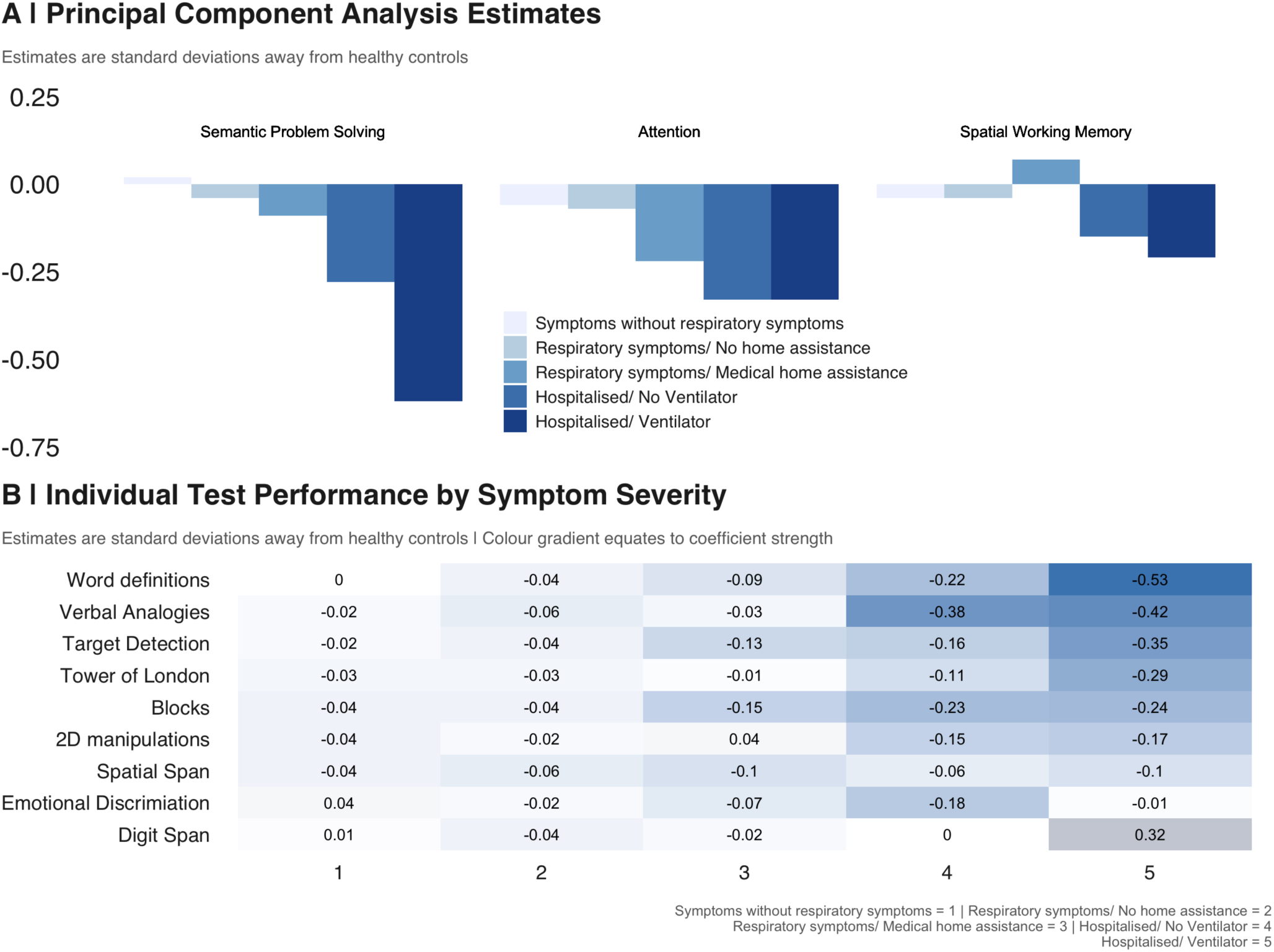
Domain sensitivity of COVID-19 related cognitive deficits. **A |** The effect size of cognitive deficits varied across three cognitive domains, which were estimated by applying principal component analysis with varimax rotation to the nine test summary scores. Semantic problem solving was particularly reduced for people who had been put on a ventilator, but also showed a significant scaled reduction for other people who required a hospital visit. Visual Attention showed similar scaled reductions in performance for all groups who required medical assistance. Spatial working memory appeared not to be significantly affected. **B |** The scale (SD units) of the cognitive deficit varied substantially across the nine tests.

## Discussion

Our analyses provide converging evidence to support the hypothesis that COVID-19 infection likely has consequences for cognitive function that persist into the recovery phase. The observed deficits varied in scale with respiratory symptom severity, related to positive biological verification of having had the virus even amongst milder cases, could not be explained by differences in age, education or other demographic and socioeconomic variables, remained in those who had no other residual symptoms and was of greater scale than common pre-existing conditions that are associated with virus susceptibility and cognitive problems.

The scale of the observed deficits was not insubstantial; the 0.57 SD global composite score reduction for the hospitalised with ventilator sub-group was equivalent to the average 10-year decline in global performance between the ages of 20 to 70 within this dataset. It was larger than the mean deficit of 512 people who indicated they had previously suffered a stroke (−0.40SDs) and the 1016 who reported learning disabilities (−0.49SDs). For comparison, in a classic intelligence test, 0.57 SDs equates to an 8.5-point difference in IQ.

At a finer grain, the deficits were broad, affecting multiple cognitive domains. They also were more pronounced for tests that assessed semantic problem solving and visual selective attention whilst sparing tests of simpler functions such as emotional processing and working-memory span. Notably, this profile cannot be explained by differences in the general sensitivity of our tests; e.g., Spatial Span and Digit Span scores show robust age-related differences and sensitivity to some other neurologic conditions. Instead, people who have recovered from COVID-19 infection show particularly pronounced problems in multiple aspects higher cognitive or ‘executive’ function, an observation that accords with preliminary reports of executive dysfunction in some patients at hospital discharge^14^, as well as previous studies of ventilated patients with acute respiratory distress syndrome pre-pandemic^19^.

Previous studies in hospitalised patients with respiratory disease not only demonstrate cognitive deficits, but suggest these remain for some at a 5 year follow-up ^19^. Consequently, the observation of post-infection deficits in the subgroup who were put on a ventilator was not surprising. Conversely, the deficits in cases who were not put on a ventilator, particularly those who remained at home, was unexpected. Although these deficits were on average of small scale for those who remained at home, they were more substantial for people who had received positive confirmation of COVID-19 infection. One possibility is that these deficits in milder cases may reflect the lower grade consequences of less severe hypoxia. However, as noted in the introduction, there have been case reports of other forms of neurological damage in COVID-19 survivors, including some for whom such damage was the first detected symptom^4^. Accordingly, in the current study, bio-positive cases who reported being ill with no breathing difficulties showed a 0.32SD magnitude cognitive deficit. Based on this, we propose that a timely challenge is to cross-relate the multi-dimensional profile of cognitive deficits observed here to imaging markers that can confirm and differentiate the underlying neuropathologies of COVID-19. Indeed, some of the tests reported here now are being applied alongside imaging in people recovering from severe illness with COVID-19 for that purpose.

An important consideration for any cross-group study is biased sampling. Crucially, our study promotional material did not mention COVID-19. Instead, we raised the profile via a BBC2 Horizon documentary plus news features stating that people could undertake a free online assessment to identify their greatest cognitive strengths. This mitigated biased recruitment of people who suspected that COVID-19 had affected their cognitive faculties. Including the questionnaire post assessment also mitigated the potential for questionnaire items to bias expectations of poor self-performance due to COVID-19.

Normal limitations pertaining to inferences about cause and effect from cross-sectional studies apply^3,20^. One might posit that people with lower cognitive ability have higher risk of catching the virus. We consider such a relationship plausible; however, it would not explain why the observed deficits varied in scale with respiratory symptom severity. We also note that the large and socioeconomically diverse nature of the cohort enabled us to include many potentially confounding variables in our analysis. Nonetheless, we emphasise that longitudinal research, including follow-up of this cohort, is required to further confirm the cognitive impact of COVID-19 infection and determine deficit longevity as a function of respiratory symptom severity, and other symptoms. It also is plausible that cognitive deficits associated with COVID-19 are no different to other respiratory illnesses. The observation of significant cognitive deficit associated with positive biological verification of having had COVID-19, i.e., relative to suspected COVID-19, goes some way to mitigate this possibility. Further work is required to interrelate the deficits to underlying neurological changes, and to disambiguate the associated pathological processes and cross compare to other respiratory viruses. A fuller understanding of the marked deficits that our study shows will enable better preparedness in the post-pandemic recovery challenges.

## Materials and Methods

### Study promotion

The Great British Intelligence Test is an ongoing collaborative citizen science project with BBC2 Horizon that launched in late December 2019. At the beginning of January, articles promoting the study were placed on the Horizon homepage, BBC News homepage and main BBC homepage, and circulated via news meta-apps. They remained in prominent positions within the public eye throughout January. In May, aligned with report of initial results considered of interest to the general public via a BBC2 Horizon documentary, there was a further promotional push. This led to high recruitment in the months of January and May, with lower, but still substantial recruitment between and after these dates.

### Data collection

The study was promoted as a free way for people to test themselves in order to find out what their greatest personal cognitive strengths were. It comprised a sequence of nine tests from the broader library that is available via the Cognitron server based on prior data showing that they can be used to measure distinct aspects of human cognition, spanning planning/reasoning, working memory, attention and emotion processing abilities, in a manner that is sensitive to population variables of interest whilst being robust against the type of device that a person is tested on. In this respect, the battery of tests should not be considered an IQ test in the classic sense, but instead, is intended to differentiate aspects of cognitive ability on a finer grain. The tests also had been optimised for application with older adults and people with mild cognitive and motor impairments.

All Cognitron tests were programmed in html5 with JavaScript by AH and WT. They were hosted on a custom server system on the Amazon EC2 that can support diverse studies via custom websites. The server system was specifically developed to handle spikey acquisition profiles that are characteristic of main-stream media collaborative studies, fitting the number of server instances in an automated manner to rapid changes in demand. Here, maximum concurrent participants landing on the website information page was >36,000, with this occurring at the point of the documentary airing on BBC2 in May.

After the nine cognitive tests, participants were presented with a detailed questionnaire with items capturing a broad range of socio-demographic, economic, vocational and lifestyle variables. During May, in response to the COVID-19 pandemic, the questionnaire was extended to include items pertaining to the direct and indirect impact of the virus, along with questions regarding common pre-existing medical conditions. At the time of writing, this had been completed by 84,285 adults, predominantly within the UK. People under the age of 16 were not excluded. Instead, they were presented with an abbreviated questionnaire that did not include COVID-19 related items. This decision was made to help ensure accelerated approval via the ethics board.

On completing the questionnaire, participants were provided with a summary report of their performance relative to all other people who had undertaken each of the tests, which highlighted the cognitive domains that they performed relatively highest on. This report was used as a way to motivate people to take part in the study. The ordering of events as outlined above was designed to mitigate biases. Specifically, the study did not advertise as having a COVID-19 related questionnaire, avoiding biased sampling of people who were concerned that the illness had reduced their cognitive functions. Furthermore, when filling out the questionnaire, participants were yet to be shown how their performance compared to the normative population, thereby avoiding biasing the questionnaire responses.

### Data pre-processing

All processing and analysis steps were conducted in MATLAB by AH with assistance from WT. Visualisation was conducted in R (v4.0.2) by JMB. Pre-processing steps were as follows. Participants under 16 or who had not completed the extended questionnaire were removed from the analysis. Each test was designed to produce one primary accuracy-based performance measure (details of test designs are provided below). Values more than 5 standard deviations from the mean were winsorised. Nuisance variables were factored out by applying a generalised linear model and taking the standardised residuals forwards for analysis relative to the variables of interest. This two-step approach was chosen because it leverages the very large data when taking into account broadly applicable nuisance variables such as age whilst ensuring that the model applied to examine effects of interest had minimal possible complexity, thereby reducing any propensity for overfit when contrasting between smaller sub-groups. Nuisance variables were age, sex, racial-ethnicity, gender, handedness, first language (English vs other), country of residence (UK vs other), education level, vocational status and annual earning. Age in years was taken to the third order in the model to fit precisely the nonlinear age curves that are characteristic of the tests.

### Composite score estimation

Composite scores were extracted from the data in two steps. First, an overall composite score was estimated across all nine tests by extracting the first eigenvector. Then, principal component analysis was conducted with varimax rotation. We conformed to the Kaiser convention of including components that had eigenvalues > 1, producing 3 orthogonal components. Examination of the rotated component loading matrix produced the expected easily interpretable solution. Specifically, the first component was labelled ‘Semantic Problem Solving’ as the heaviest loadings for it were Rare Word Definitions, which involves assigning definitions to rare words, and Analogical Reasoning, which requires the mapping of rules and relationships between different semantic contexts. The second component was labelled ‘Visual Attention’ as the heaviest weightings were for 2D Mental Rotations followed by Target Detection, both of which require the rapid processing of visual arrays that have complex combinations of features. The third component was labelled ‘Spatial Working Memory’ as it comprised all of the spatial tests, including the Spatial Span, which measures working memory capacity, and Tower of London and Block Rearrange, which involve spatial problem solving.

### Linear models

The overall summary score, three component scores and nine individual test scores with nuisance variables factored out, were taken forwards for analysis with general linear modelling. The first analysis examined differences in scores relative to people who were not ill for those who reported that they believed they had recovered from the COVID-19 illness. These were subdivided along an approximate severity scale into (i) those who did not have trouble breathing, (ii) those who had breathing problems but received no medical assistance, (ii) those who had breathing problems and received medical assistance at home, (iv) those who were taken to hospital but were not put on a ventilator and (v) those who were fitted with a ventilator. Further models were then run focused on the summary score to examine if the observed deficits had a basis in other factors. These included as additional factors in the GLM (i) positive confirmation of COVID-19 infection through a biological test, (ii) people who reported residual COVID-19 symptoms, (and (iii) common pre-existing medical conditions that affect the respiratory system or immune system and that are associated with cognitive deficits. Further analyses that are not reported here include (iv) pre-existing neurological conditions and (v) pre-existing psychiatric conditions. These are in preparation for a further article, but we can report that inclusion of these variables does not diminish the COVID-19 effects.

### Task designs

The cognitive tests included in this study (and three more recently added tests) can be viewed at https://gbit.cognitron.co.uk. In brief, the main study included nine tests that based on previous analyses were known to be robust across devices, sensitive to population variables of interest such as age, gender and education level, manageable for older adults and patients with mild cognitive or motor deficits, and not so strongly correlated as to measure just one overarching ability. Details of individual task designs are in supplementary figures S1-9. Questionnaire items analysed in this study are outlined in Supplementary Table S1

## Data Availability

Data available upon request

## Acknowledgements

Dr Hampshire is supported by the UK Dementia Research Institute and Biomedical Research Centre at Imperial College London with technology development supported by EU-CIG EC Marie-Curie CIG and NIHR grant II-LB-0715-20006. William Trender is supported by the EPSRC Center for Doctoral Training in Neurotechnology. Dr Chamberlain’s role in this study was funded by a Wellcome Trust Clinical Fellowship (Reference 110049/Z/15/Z). Joseph M Barnby is supported by the UK Medical Research Council (MR/N013700/1) and King’s College London member of the MRC Doctoral Training Partnership in Biomedical Sciences. Mitul Mehta is in part supported by the National Institute for Health Research (NIHR) Biomedical Research Centre at South London and Maudsley NHS Foundation Trust and King’s College London. The views expressed are those of the author(s) and not necessarily those of the NHS, the NIHR or the Department of Health and Social Care. We would like to acknowledge COST Action CA16207 ‘European Network for Problematic Usage of the Internet’, supported by COST (European Cooperation in Science and Technology); and the support of the National UK Research Network for Behavioural Addictions (NUK-BA).

## Supplementary Materials for

### Materials and Methods

#### Test designs

The cognitive tests included in this study (and three more recently added tests) can be viewed at https://gbit.cognitron.co.uk. In brief, the main study included nine tests that based on previous analyses were known to be robust across devices, sensitive to population variables of interest such as age, gender and education level, manageable for older adults and patients with mild cognitive or motor deficits, and not so strongly correlated as to measure just one overarching ability. Designs are in figures S1-9.

---

**Figure S1.**
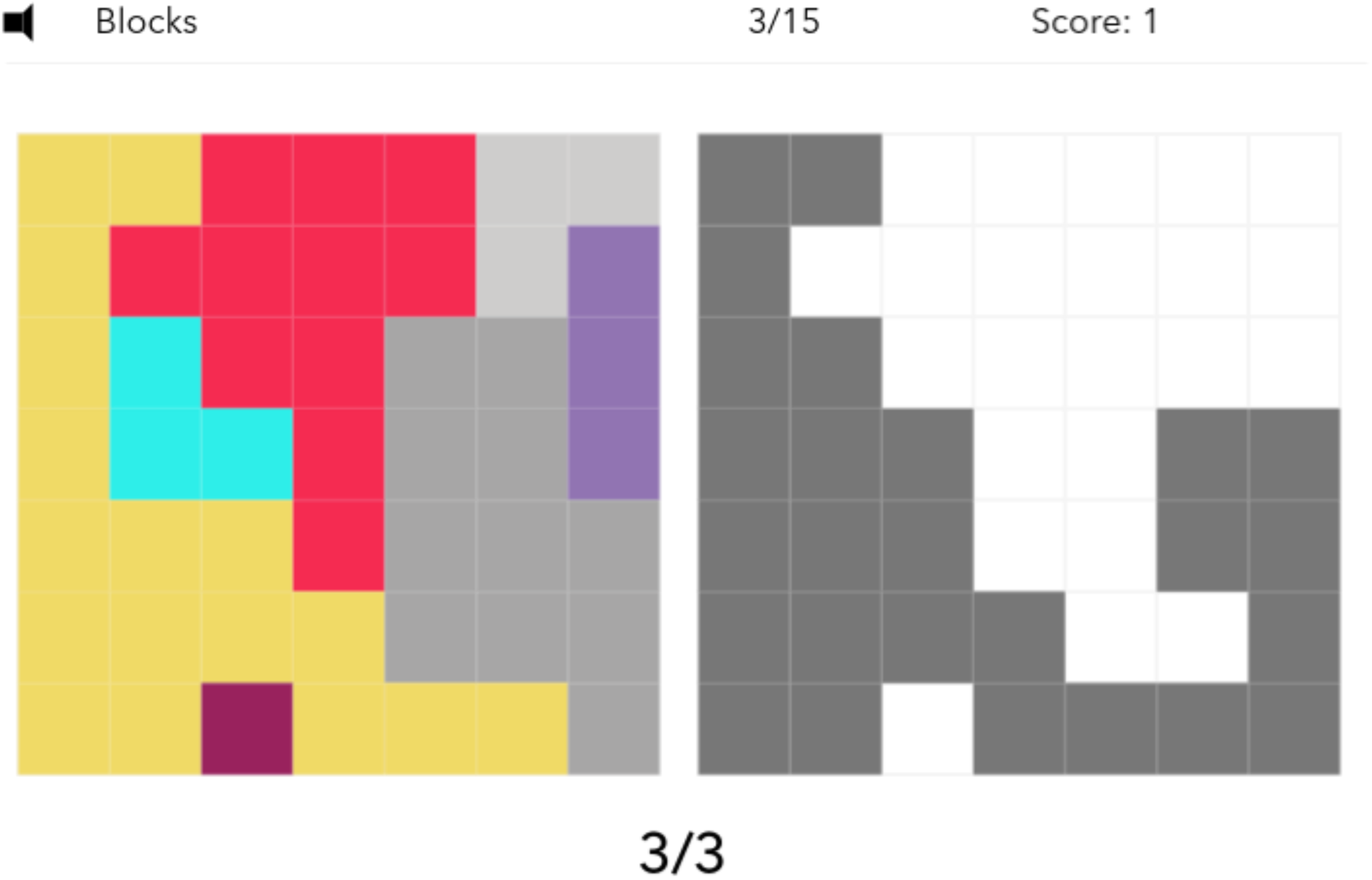
Block Rearrange. The Block Rearrange test measures spatial problem solving. The participant is presented with a grid of coloured blocks on the left-hand side of the screen and on the right-hand side, a black silhouette made up of a subset of the shapes on the left. The participant must make the shape of the left-hand blocks match the silhouette on the right-hand side by removing blocks. The blocks fall under gravity. The test comprises 15 trials of varying difficulty. The difficulty is modulated by two factors; the number of blocks needed to remove and the number of blocks that must fall in order to reach the target silhouette. Each trial is terminated if, either the target silhouette is reached (correct trial) or an incorrect block is removed (incorrect trial). The outcome measure is the total number of correct trials. Population mean= 10.9 SD = 2.93.

**Figure S2.**
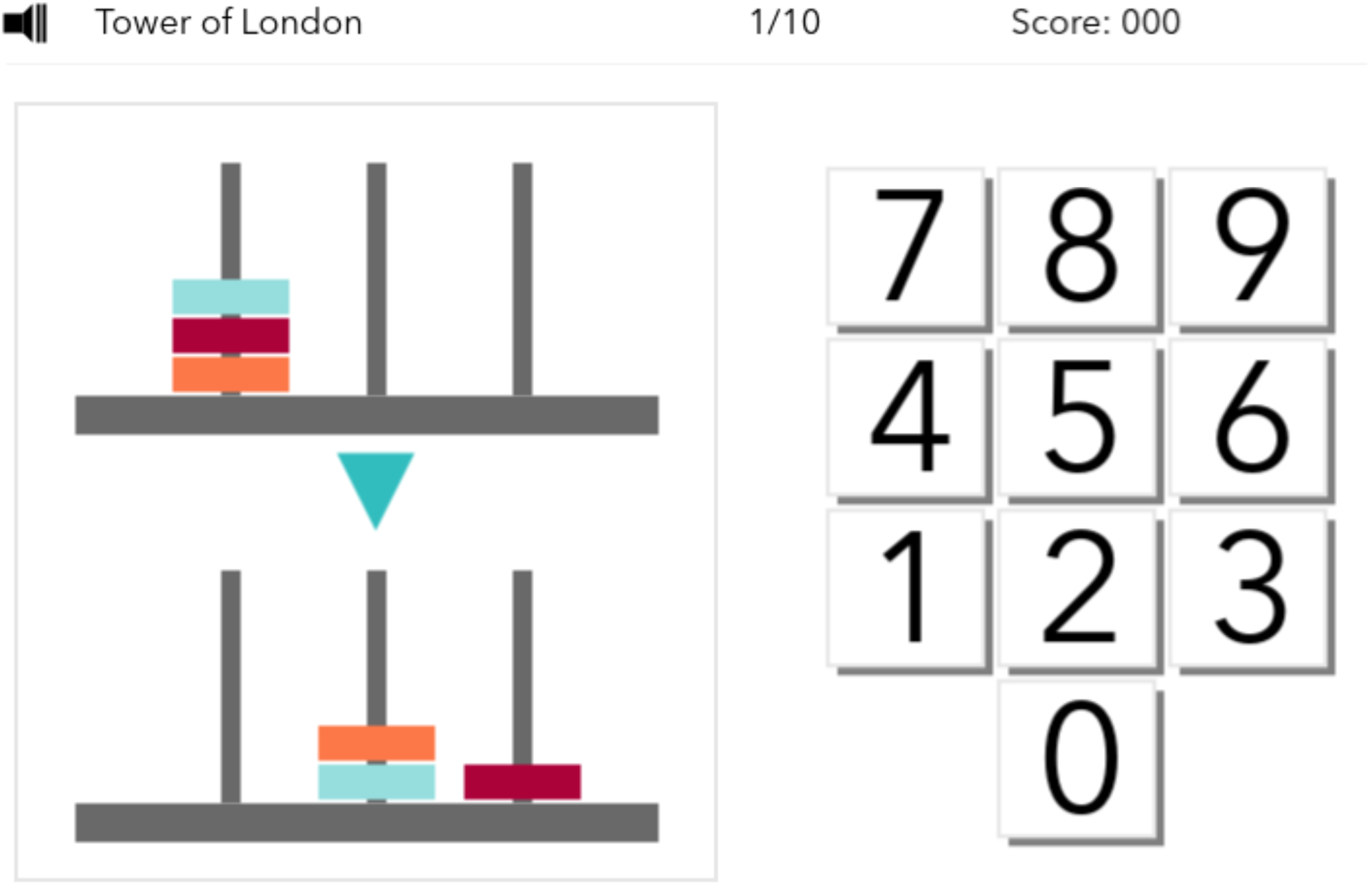
Tower of London. The Tower of London test measures spatial planning. It is a variant on the original Tower of London Test (Shallice, 1982). The participant is shown two sets of three prongs with coloured beads on them. The first set is the initial state and the second set is the target state. The participant must work out the lowest number of moves it would take to transition from the initial state to the target state. They must then input this number using an on-screen number pad. This differs from the original test in that the participant is not allowed to move the beads, all calculation and planning must be done in their head. This is to prevent correct answers being reached through iterative error correction. The test consists of 10 trials of variable difficulty. The difficulty is scaled using the number of beads and the convolutedness, defined as the number of moves that must be made that do not place a bead in its final target position. The outcome measure is the total number of correct trials. Population mean= 6.57, SD = 2.62.

**Figure S3.**
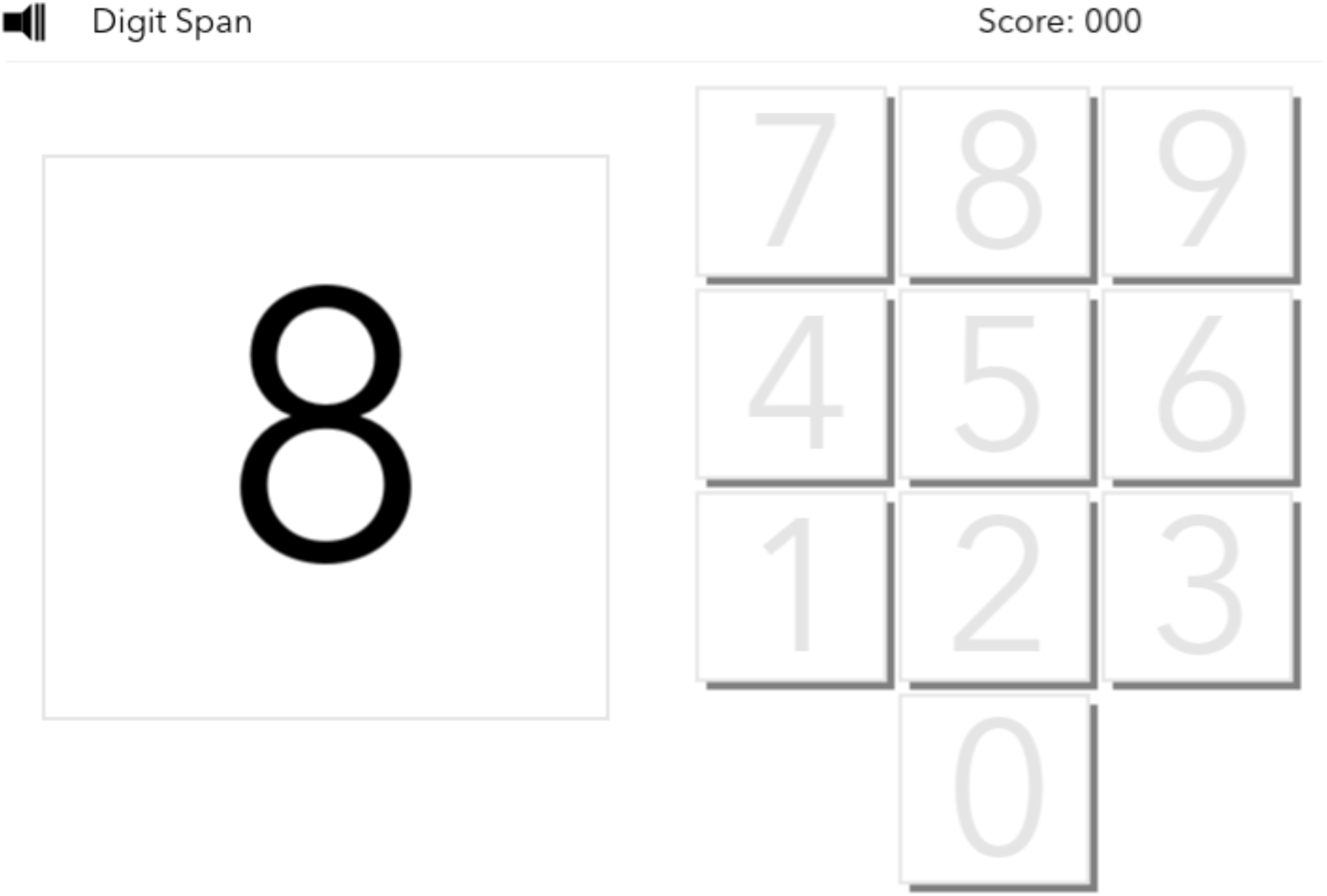
Digit Span. Digit Span is a computerised variant on the verbal working memory component of the WAIS-R intelligence test (Weschler, 1981). Participants view a sequence of digits that appear on the screen one after another. Subsequently, they repeat the sequence of numbers by entering them using an on-screen number pad. The difficulty is incremented using a ratchet system, every time a sequence is recalled correctly, the length of the subsequent sequence is incremented by one. The test is terminated when three consecutive mistakes are made on a particular sequence length. The outcome measure is the maximum sequence length correctly recalled. Minimum level = 2, maximum level = 20, ISI = 0ms, encoding time = 1000ms. Population mean = 6.98, SD = 1.58.

**Figure S4.**
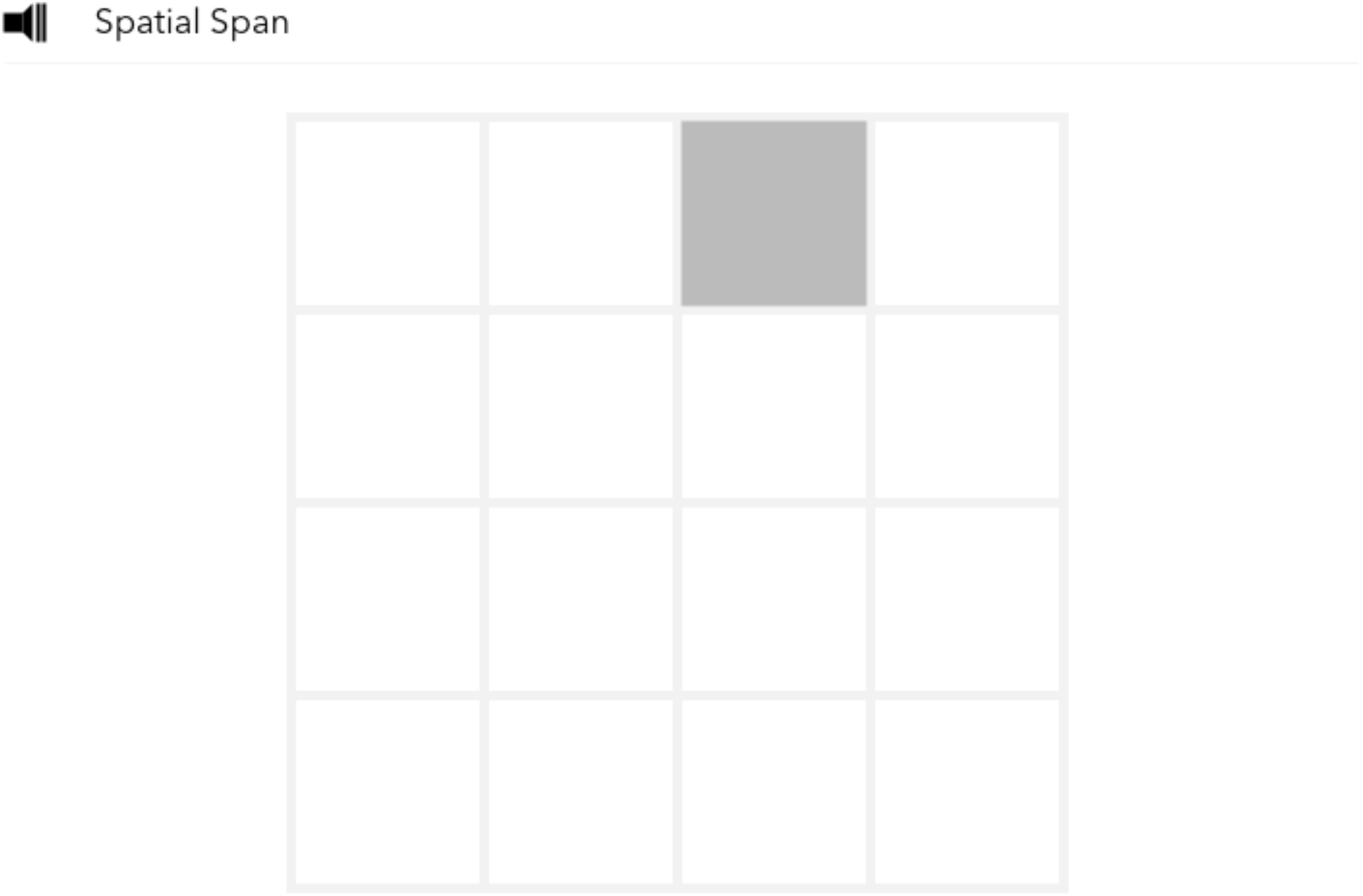
Spatial Span. The Spatial Span test measures spatial short-term memory capacity. It is a variant on the classic Corsi Block Tapping Test (Corsi, 1972). The participant is presented with a 4 x 4 grid, onto which is displayed a sequence of squares in different positions in the grid. The participant must then click the squares in the order that they were highlighted. The difficulty is incremented using a ratchet system, every time a sequence is recalled correctly, the length of the subsequent sequence is incremented by one. The test is terminated when three consecutive mistakes are made on a particular sequence length. The outcome measure is the maximum sequence length correctly recalled. Minimum level = 2, maximum level = 16, ISI = 0ms, encoding time = 1500ms. Population mean = 6.10, SD = 1.23.

**Figure S5.**
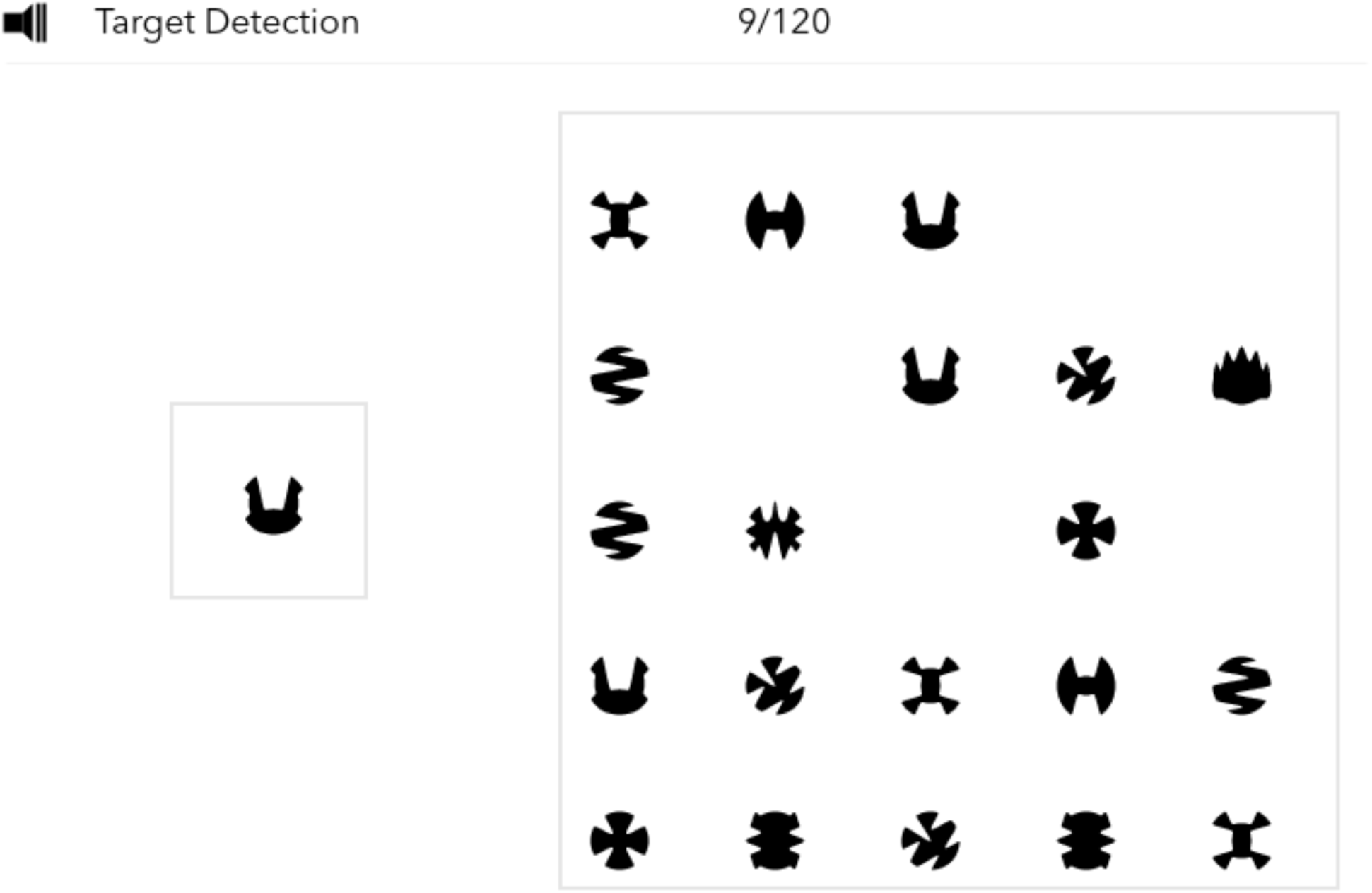
Target Detection. The target detection test measures spatial visual attention. The participant is presented with a target shape on the left of the screen and a probe area on the right side of the screen. After 3000ms, the probe area begins to fill with shapes, the participant must identify and click the target shape while ignoring the distractor shapes. Shapes are added every 1000ms and a subset of the shapes in the probe area are removed every 1000ms. The trial runs for a total of 120 addition/removal cycles. The target shape is included in the added shapes pseudo randomly, at a frequency of 12 in 20 cycles. The outcome measure is the total number of target shapes clicked. Population mean = 57.2, SD = 11.8.

**Figure S6.**
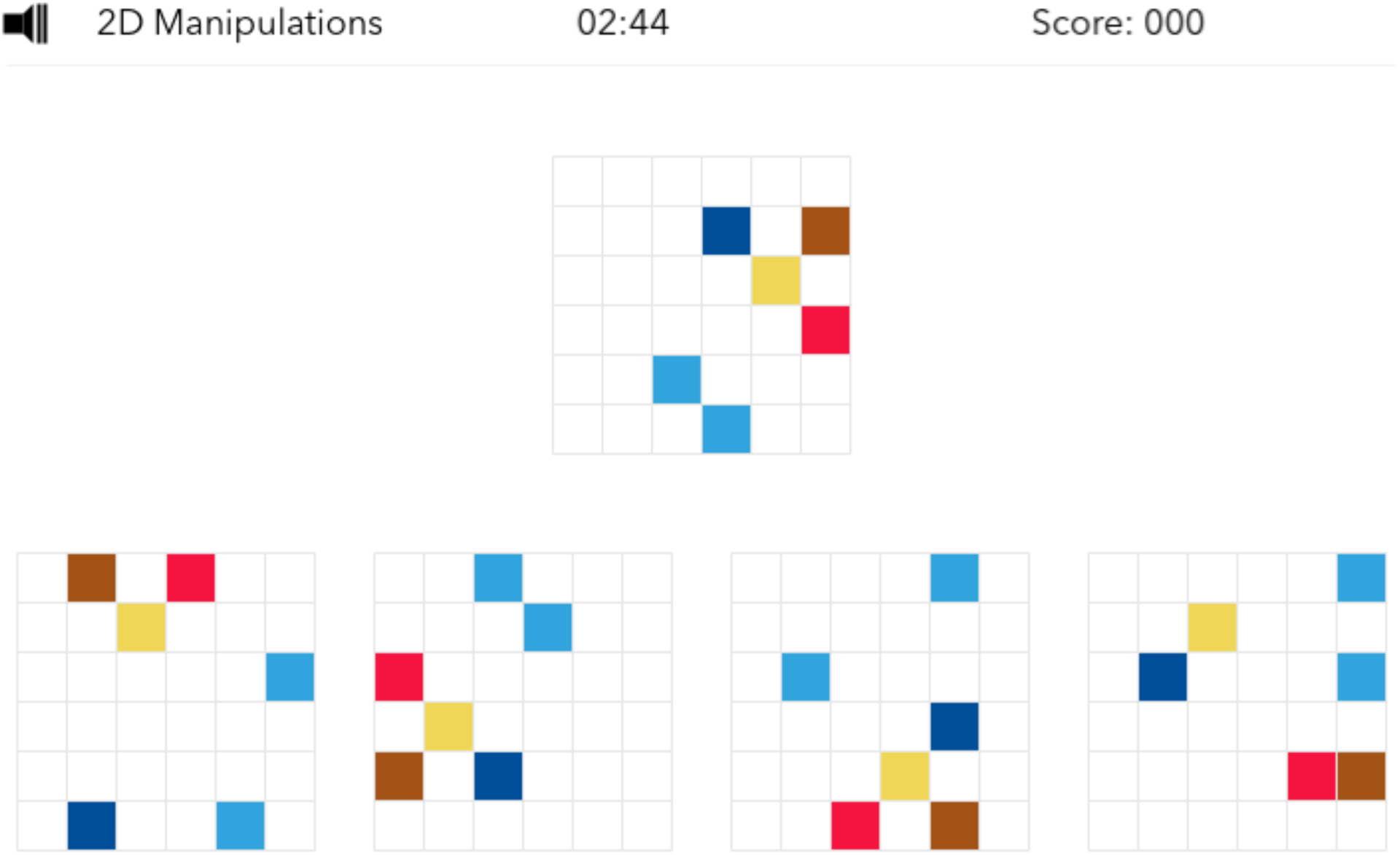
2D Mental Rotation. The 2D Mental rotation test measures the ability to spatially manipulate objects in mind (Silverman et al., 2000). In this version of the test, a grid with coloured squares is presented at the top of the screen, with a further four grids with coloured squares presented below (i.e. probe grids). One of the four grids is identical to the target grid above but is rotated by either 90, 180 or 270 degrees whilst the other grids differ by five squares. To obtain maximum points, the participant must indicate which of the four grids is identical to target, solving as many problems as possible within three minutes. For every correct response, the total score increases by one. The outcome measure is the total score. Population mean = 26.8, SD = 8.35.

**Figure S7.**
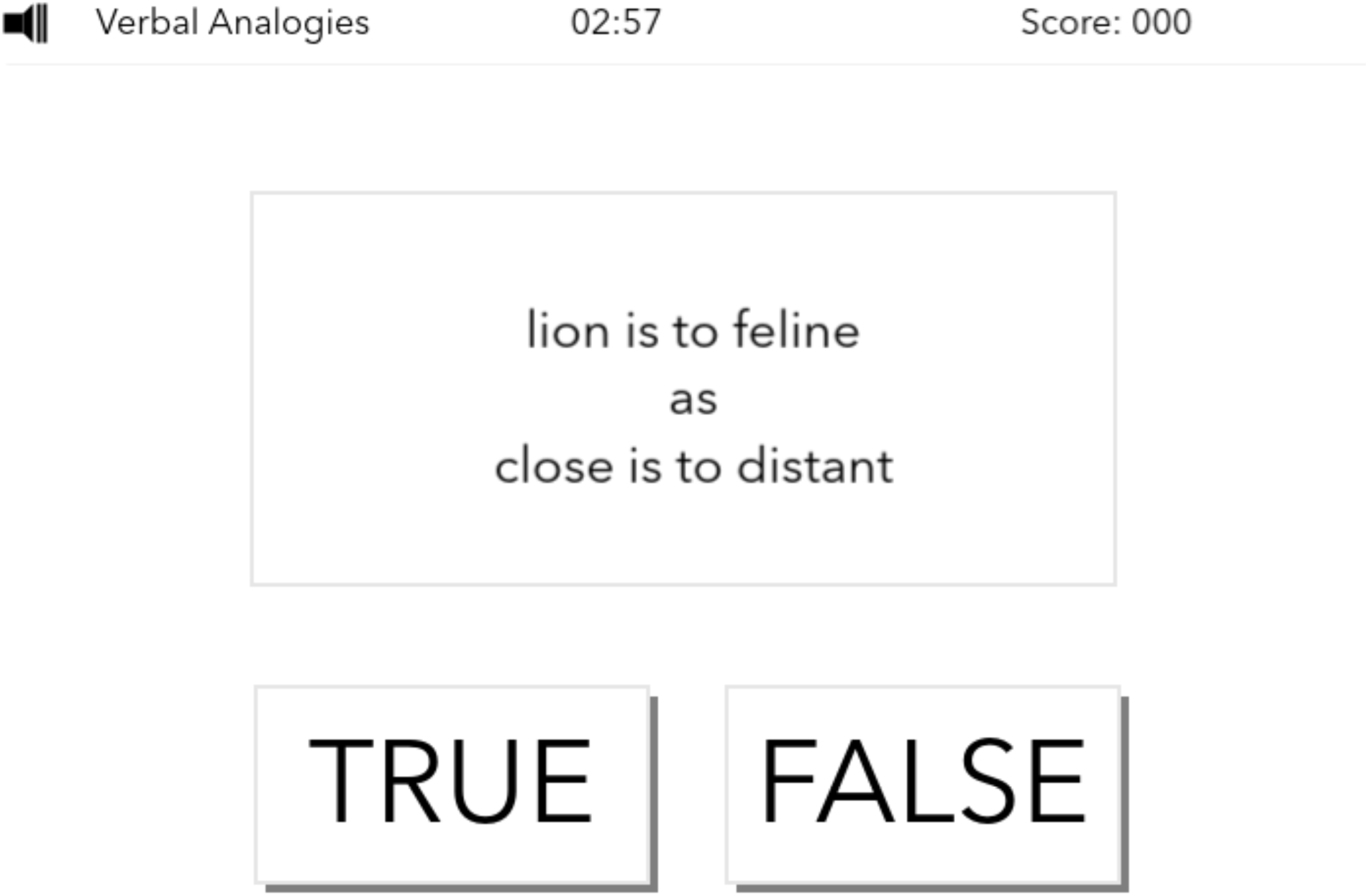
Analogical Reasoning. The Analogical Reasoning test measures semantic reasoning abilities. In this version of the test, participants are presented with two written relationships that they must decide have the same type of association or not (e.g. “Lion is to feline as cabbage is to vegetable”). Participants must indicate their decision by selecting the True or False buttons presented below the written analogies. Analogies are varied across semantic distance to modulate difficulty and associations types switch throughout the sequence of trials. To obtain maximum points, participants must solve as many problems as possible within three minutes. For every correct response, the total score increases by one. For every incorrect response, the total score decreases by one. The outcome measure is the total score.Population mean = 24.1, SD = 11.5.

**Figure S8.**
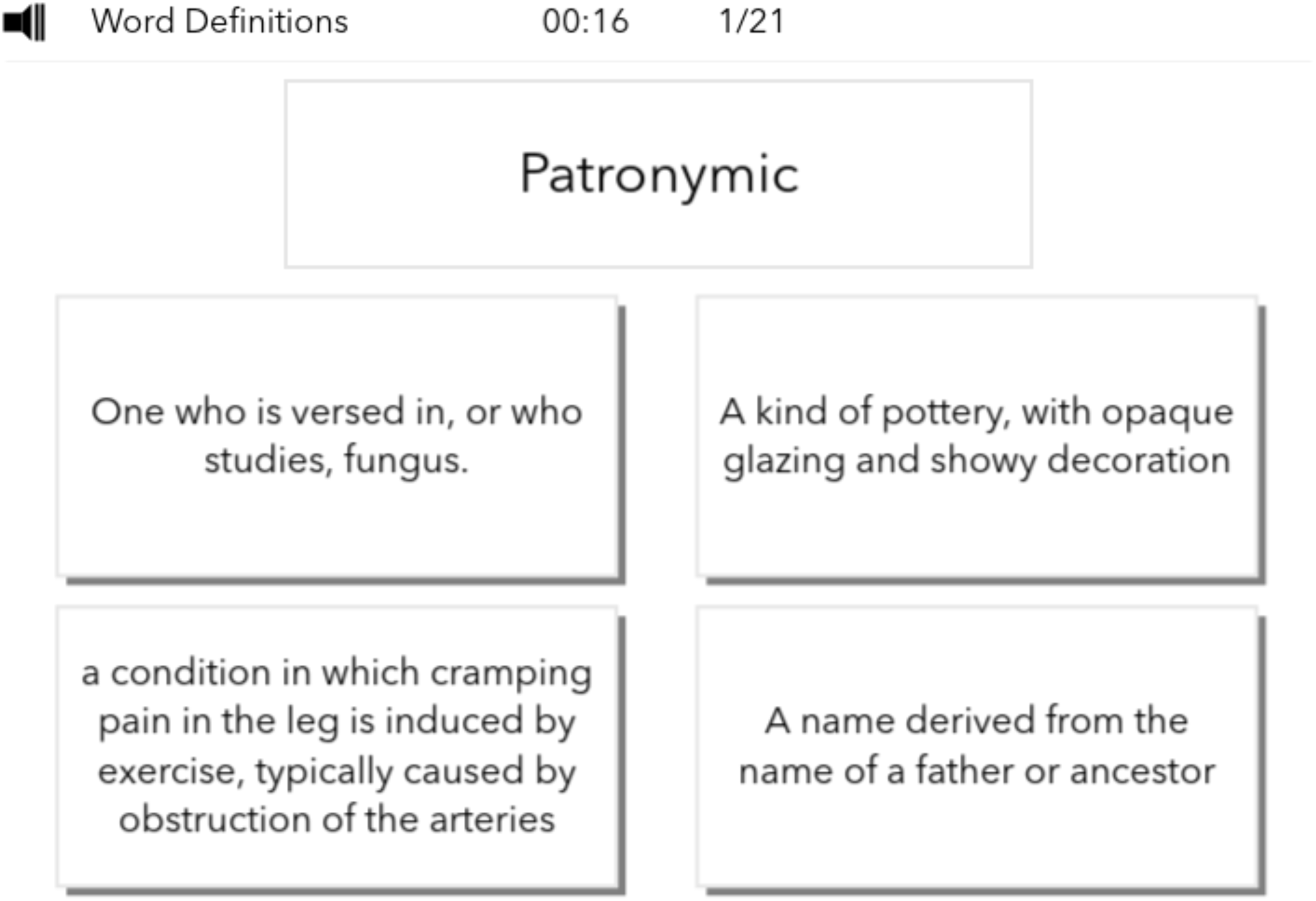
Rare Word Definitions. In this test, individuals are assessed on their ability to identify the correct definitions of a words. Participants are presented with a word accompanied by four descriptive statements. They must decide which of the four statements provides the correct definition of the word. Words vary based on their frequency of use in English written language, resulting in rare and commonly used words to be presented. For each word, the participant has twenty seconds to choose a definition. To obtain maximum points, participants must answer 21 word-definitions correctly. For every correct response, the total score increases by one point. The outcome measure is the total score. Population mean = 16.7, SD = 2.84.

**Figure S9.**
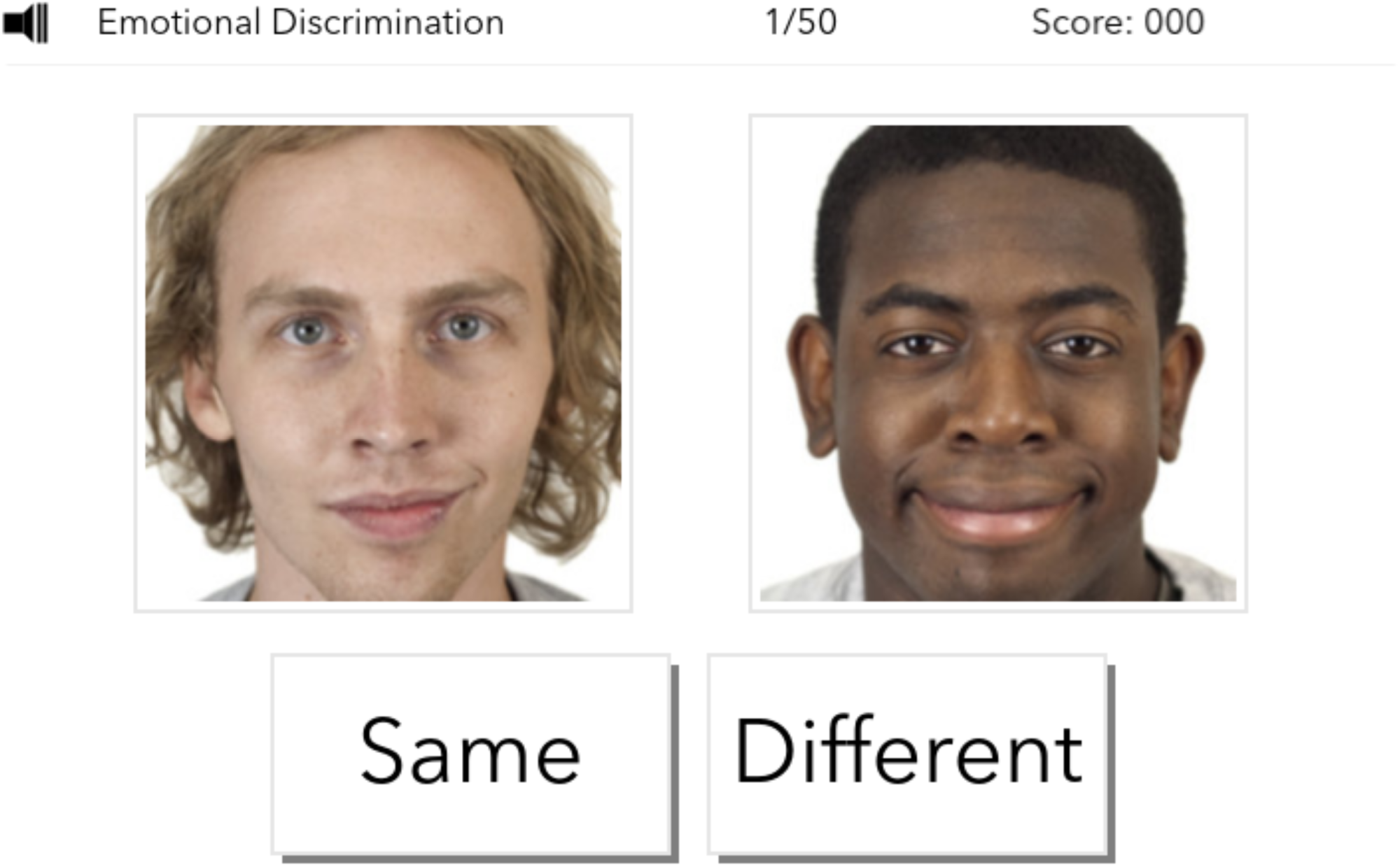
Face Emotional Discrimination. This test measures an individual’s ability to identify and discern between emotions. Participants are presented with pictures of two people, each expressing a particular emotion (e.g. happy, neutral, angry, scared). They must decide if the emotions expressed by each person are the same or different. Trials vary based on the emotions used as well as whether individuals have congruent vs. incongruent emotional expressions. To obtain maximum points, participants must complete 50 trials as accurately as possible. For every correct answer, the total score increases by one point. The outcome measure is total score.Population mean = 42.6, SD = 3.50.

**Table S1.**
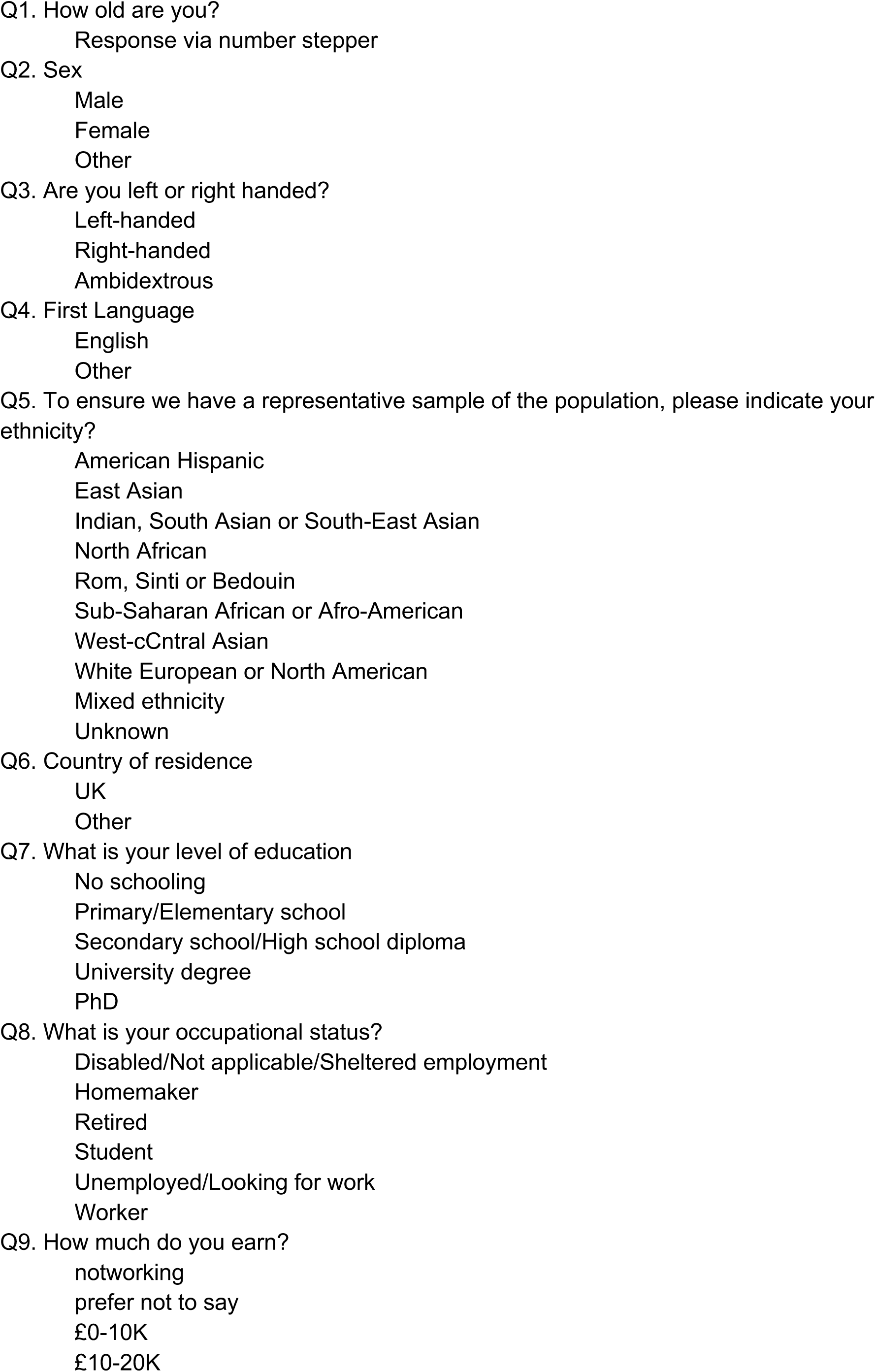
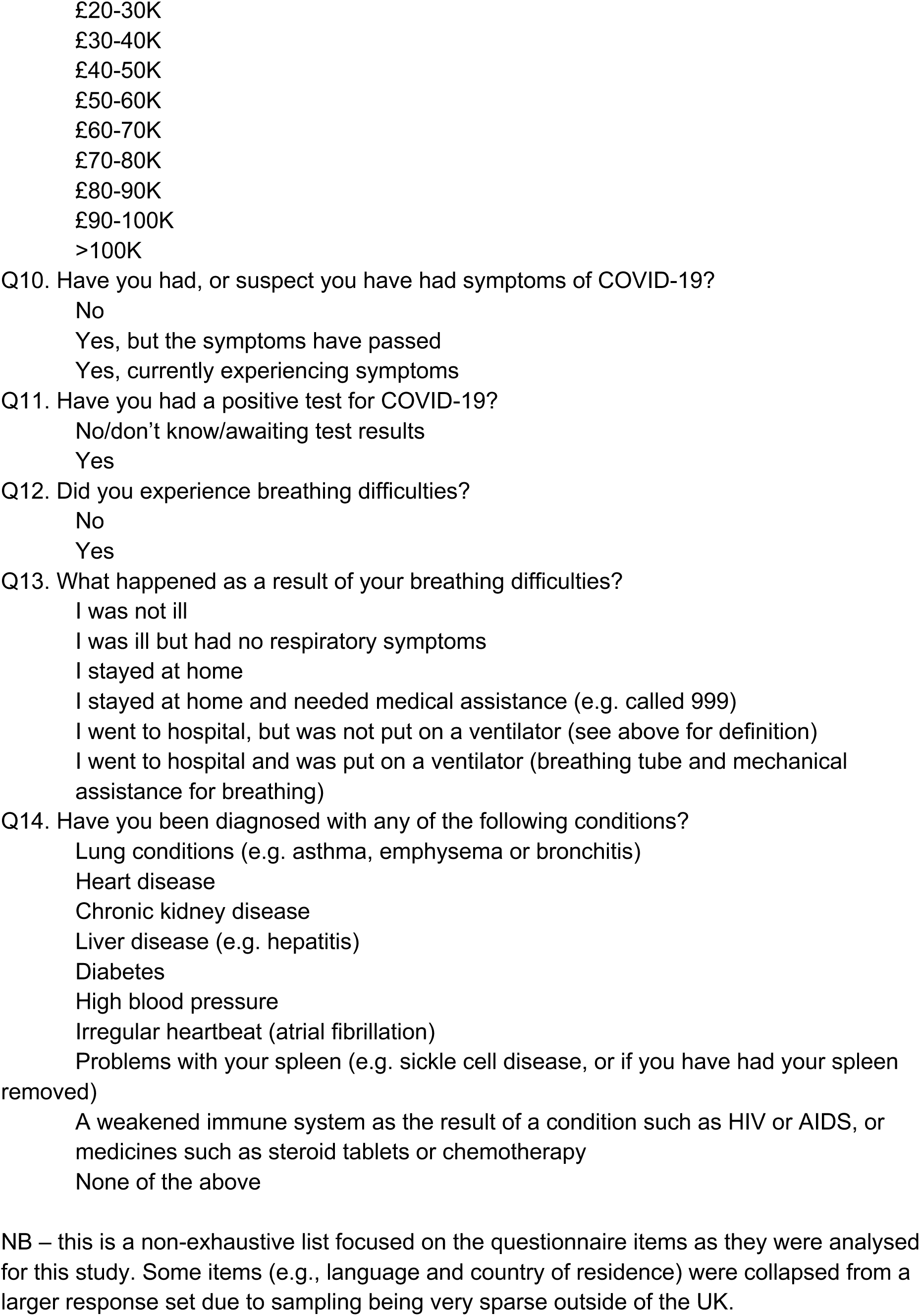
Questionnaire items analysed in this study

**Table S2a.** age distribution and severity counts per age

|  | Was not ill | Ill but had no respiratory symptoms | Respiratory symptoms, no assistance at home | Respiratory symptoms, medical assistance at home | Went to hospital but was not put on a ventilator | Went to hospital and was put on a ventilator |
| --- | --- | --- | --- | --- | --- | --- |
| **16** | 783 | 132 | 29 | 0 | 0 | 2 |
| **17** | 667 | 99 | 30 | 0 | 1 | 0 |
| **18** | 680 | 111 | 22 | 0 | 4 | 1 |
| **19** | 657 | 101 | 37 | 1 | 1 | 0 |
| **20** | 645 | 117 | 38 | 5 | 0 | 2 |
| **21** | 696 | 101 | 41 | 0 | 0 | 3 |
| **22** | 721 | 147 | 34 | 1 | 3 | 4 |
| **23** | 860 | 121 | 41 | 2 | 1 | 1 |
| **24** | 833 | 124 | 39 | 3 | 1 | 1 |
| **25** | 961 | 154 | 62 | 4 | 1 | 0 |
| **26** | 1047 | 176 | 55 | 2 | 1 | 2 |
| **27** | 1101 | 190 | 86 | 3 | 4 | 2 |
| **28** | 1195 | 178 | 85 | 3 | 2 | 1 |
| **29** | 1215 | 209 | 72 | 5 | 2 | 1 |
| **30** | 1269 | 193 | 73 | 1 | 2 | 3 |
| **31** | 1186 | 176 | 80 | 7 | 5 | 1 |
| **32** | 1272 | 175 | 82 | 2 | 4 | 1 |
| **33** | 1151 | 171 | 67 | 4 | 4 | 0 |
| **34** | 1058 | 171 | 80 | 3 | 2 | 1 |
| **35** | 1046 | 172 | 74 | 6 | 1 | 1 |
| **36** | 1160 | 153 | 69 | 3 | 4 | 2 |
| **37** | 1009 | 180 | 63 | 3 | 3 | 1 |
| **38** | 1105 | 194 | 84 | 6 | 2 | 0 |
| **39** | 1321 | 204 | 97 | 4 | 5 | 0 |
| **40** | 1280 | 211 | 77 | 4 | 6 | 4 |
| **41** | 1191 | 185 | 82 | 6 | 5 | 0 |
| **42** | 1231 | 198 | 59 | 4 | 2 | 1 |
| **43** | 1181 | 179 | 71 | 4 | 1 | 1 |
| **44** | 1231 | 148 | 74 | 5 | 2 | 2 |
| **45** | 1427 | 218 | 82 | 7 | 1 | 1 |
| **46** | 1338 | 220 | 92 | 8 | 3 | 1 |
| **47** | 1413 | 207 | 69 | 11 | 3 | 0 |
| **48** | 1518 | 180 | 92 | 10 | 3 | 3 |
| **49** | 1610 | 215 | 78 | 2 | 4 | 1 |
| **50** | 1633 | 239 | 87 | 8 | 5 | 0 |
| 51 | 1458 | 193 | 91 | 8 | 3 | 3 |
| 52 | 1554 | 201 | 81 | 1 | 6 | 0 |
| 53 | 1583 | 212 | 86 | 5 | 6 | 1 |
| 54 | 1672 | 196 | 82 | 1 | 4 | 3 |
| 55 | 1723 | 212 | 90 | 3 | 1 | 2 |
| 56 | 1606 | 183 | 68 | 3 | 6 | 1 |
| 57 | 1592 | 175 | 73 | 2 | 2 | 0 |
| 58 | 1558 | 184 | 68 | 3 | 3 | 0 |
| 59 | 1550 | 181 | 58 | 1 | 7 | 0 |
| 60 | 1575 | 175 | 62 | 0 | 1 | 0 |
| 61 | 1460 | 121 | 48 | 1 | 3 | 0 |
| 62 | 1455 | 155 | 46 | 0 | 1 | 0 |
| 63 | 1423 | 144 | 43 | 1 | 3 | 1 |
| 64 | 1367 | 132 | 40 | 1 | 0 | 1 |
| 65 | 1294 | 142 | 45 | 1 | 2 | 1 |
| 66 | 1221 | 100 | 28 | 0 | 1 | 0 |
| 67 | 1161 | 91 | 22 | 2 | 2 | 0 |
| 68 | 1010 | 74 | 32 | 0 | 0 | 0 |
| 69 | 901 | 60 | 21 | 2 | 1 | 0 |
| 70 | 890 | 65 | 17 | 2 | 2 | 0 |
| 71 | 739 | 50 | 9 | 0 | 0 | 0 |
| 72 | 798 | 36 | 10 | 1 | 0 | 0 |
| 73 | 679 | 48 | 11 | 0 | 0 | 1 |
| 74 | 398 | 23 | 11 | 0 | 0 | 0 |
| 75 | 379 | 24 | 5 | 0 | 1 | 0 |
| 76 | 295 | 10 | 2 | 0 | 1 | 0 |
| 77 | 243 | 16 | 2 | 1 | 1 | 0 |
| 78 | 182 | 5 | 3 | 0 | 0 | 0 |
| 79 | 161 | 11 | 0 | 0 | 0 | 0 |
| 80 | 127 | 6 | 4 | 0 | 0 | 0 |
| 81 | 97 | 2 | 1 | 0 | 0 | 0 |
| 82 | 100 | 8 | 1 | 0 | 2 | 0 |
| 83 | 76 | 2 | 0 | 0 | 0 | 0 |
| 84 | 42 | 4 | 0 | 0 | 0 | 0 |
| 85 | 51 | 0 | 1 | 0 | 0 | 0 |
| 86+ | 124 | 11 | 2 | 0 | 0 | 2 |

**Table S2b.** Ethnic groups within the GBIT cohort

| Whole cohort |  | Extended questionnaire |  | Ethnicity |
| --- | --- | --- | --- | --- |
| N | % | N | % |  |
| 9599 | 2.5 | 479 | 0.6 | American Hispanic |
| 14128 | 3.7 | 891 | 1.1 | East Asian |
| 14685 | 3.9 | 2181 | 2.6 | Indian, South Asian or South-East Asian |
| 9935 | 2.6 | 1882 | 2.2 | Mixed ethnicity |
| 1091 | 0.3 | 133 | 0.2 | North African |
| 585 | 0.2 | 55 | 0.1 | Rom, Sinti or Bedouin |
| 2314 | 0.6 | 253 | 0.3 | Sub-saharan African or Afro-american |
| 5533 | 1.5 | 953 | 1.1 | Unknown |
| 1757 | 0.5 | 257 | 0.3 | West-central Asian |
| 321399 | 84.4 | 77504 | 91.6 | White European or North American |

**Table S2c.**
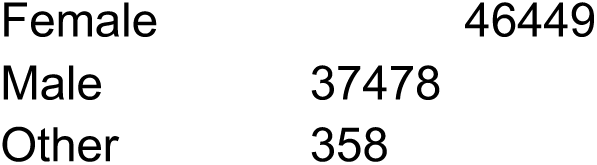
Sex

|  |  |
| --- | --- |
| Female | 46449 |
| Male | 37478 |
| Other | 358 |

**Table S2d.** Handedness

|  |  |
| --- | --- |
| Ambidextrous | 2164 |
| Left-handed | 9089 |
| Right-handed | 73032 |

**Table S2e.** First language

|  |  |
| --- | --- |
| English | 79283 |
| Other | 5002 |

**Table S2f.**
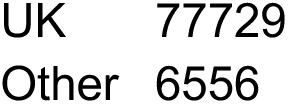
country of residence

**Table S2g.** Education level

|  |  |
| --- | --- |
| 208 | 01 No schooling |
| 1730 | 02 Primary/Elementary school |
| 30229 | 03 Secondary school/High school diploma |
| 48745 | 04 University degree |
| 3373 | 05 PhD |

**Table S2h.** Occupational status

|  |  |
| --- | --- |
| 878 | Disabled/Not applicable/Sheltered employment |
| 2727 | Homemaker |
| 16503 | Retired |
| 6487 | Student |
| 2654 | Unemployed/Looking for work |
| 54653 | Worker |
| 383 | Unknown |

**Table S2i.** Earnings

|  |  |
| --- | --- |
| 29632 | notworking |
| 1990 | prefer not to say |
| 828 | £0-10K |
| 8962 | £10-20K |
| 11423 | £20-30K |
| 10180 | £30-40K |
| 7188 | £40-50K |
| 4265 | £50-60K |
| 2514 | £60-70K |
| 1818 | £70-80K |
| 1143 | £80-90K |
| 1107 | £90-100K |
| 3235 | >100K |

**Table S3.**
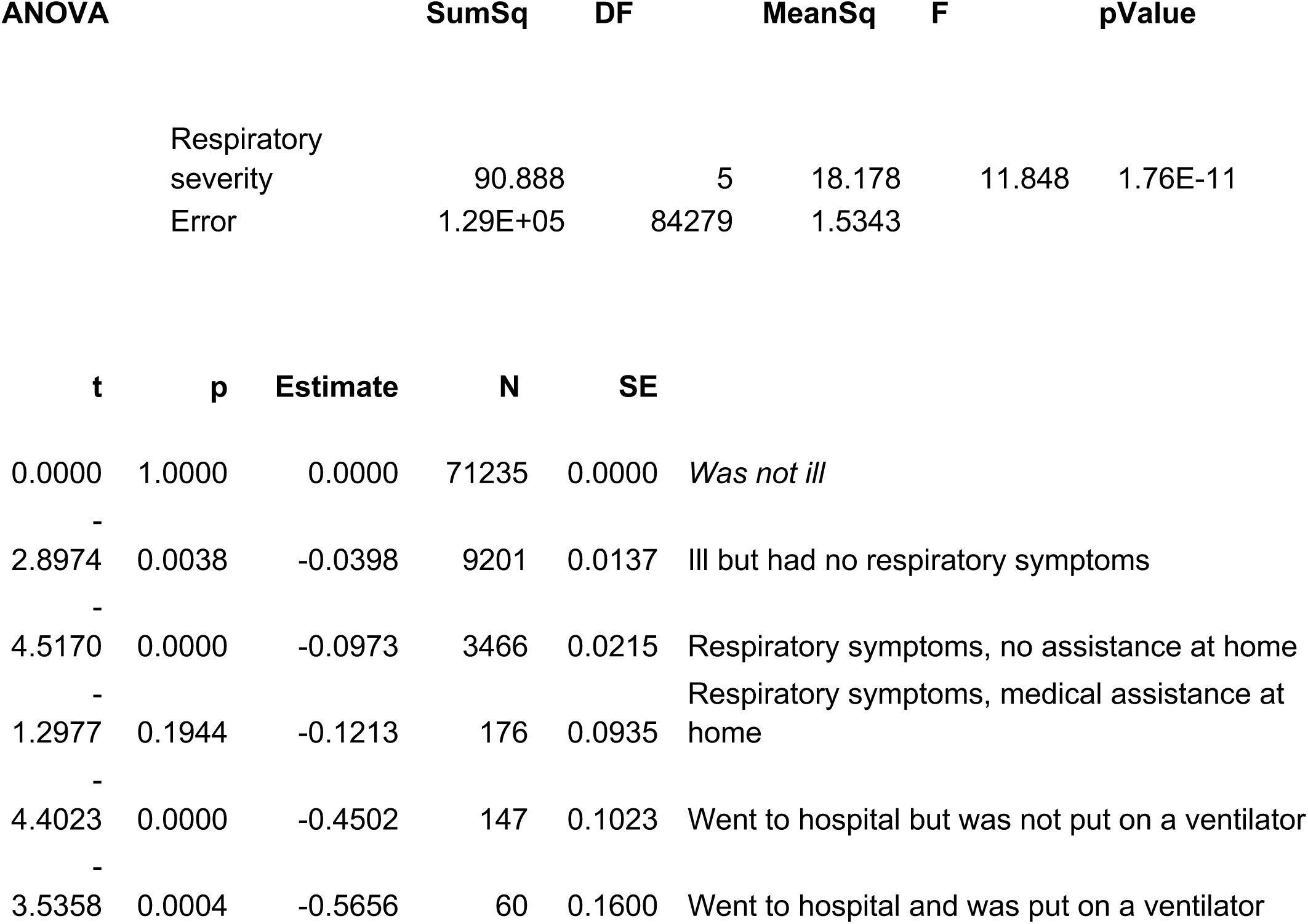
General linear model of global task performance vs. respiratory severity

**Table S4a.**
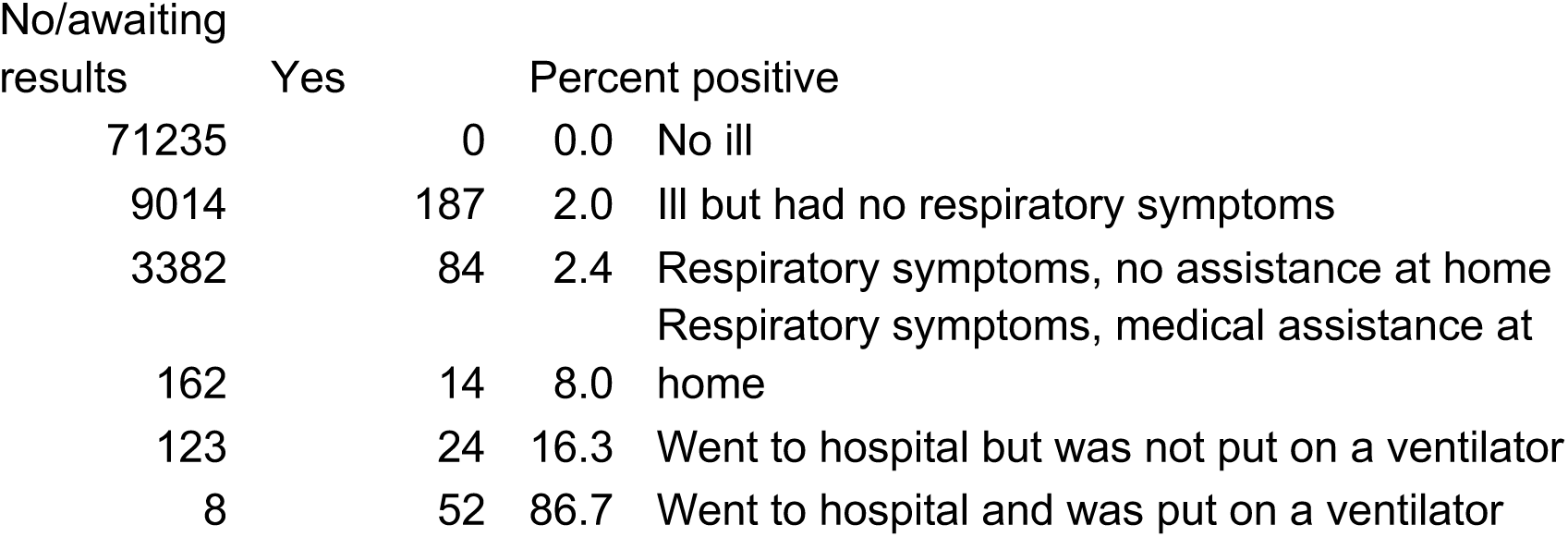
Positive COVID-19 biological test rates

**Table S4b.**
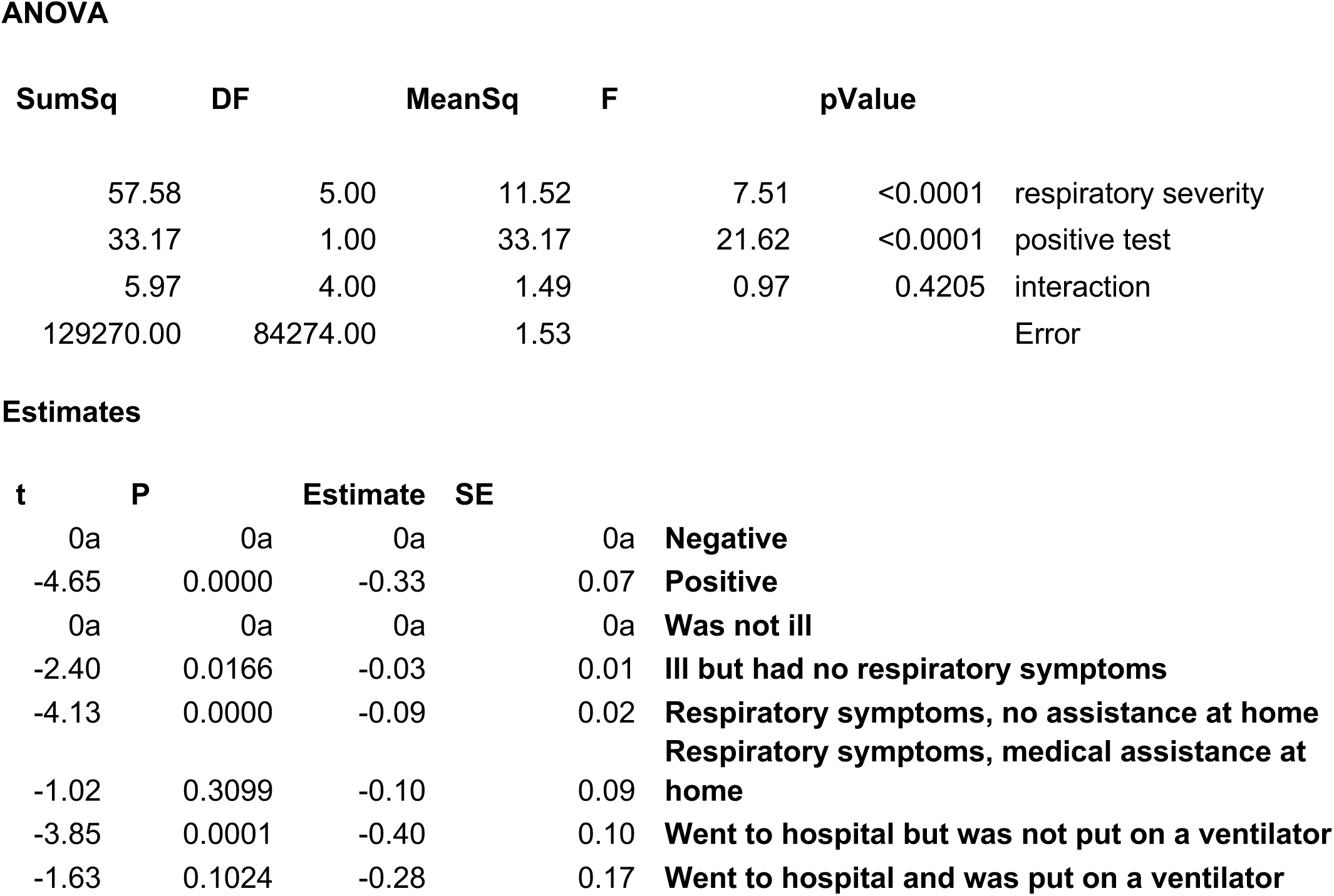
General linear model including positive COVID-19 biological test as a factor

**Table S4c.** Contrasting positive COVID-19 biological test within select groups

| Estimate | SE | tStat | pValue | N | group |
| --- | --- | --- | --- | --- | --- |
| -0.32 | 0.091 | -3.49 | 0.0005 | 187 | Ill but had no respiratory symptoms |
| -0.36 | 0.14 | -2.61 | 0.0091 | 84 | Respiratory symptoms, no assistance at home |

**Table S5a.**
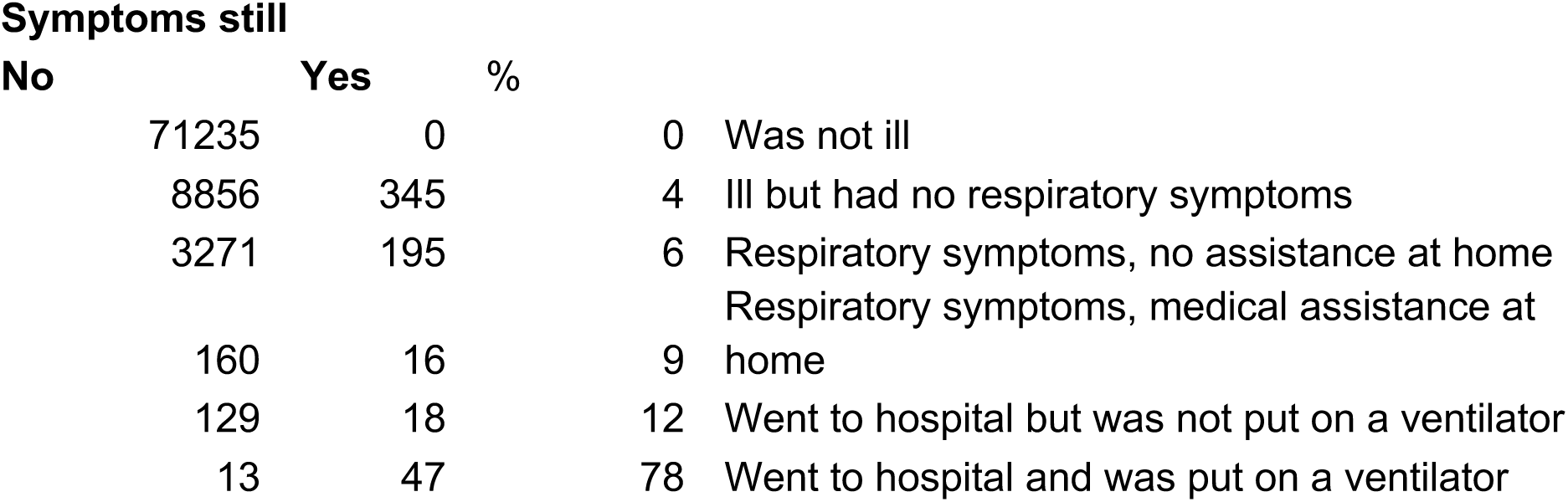
Residual symptom rates

**Table S5b.**
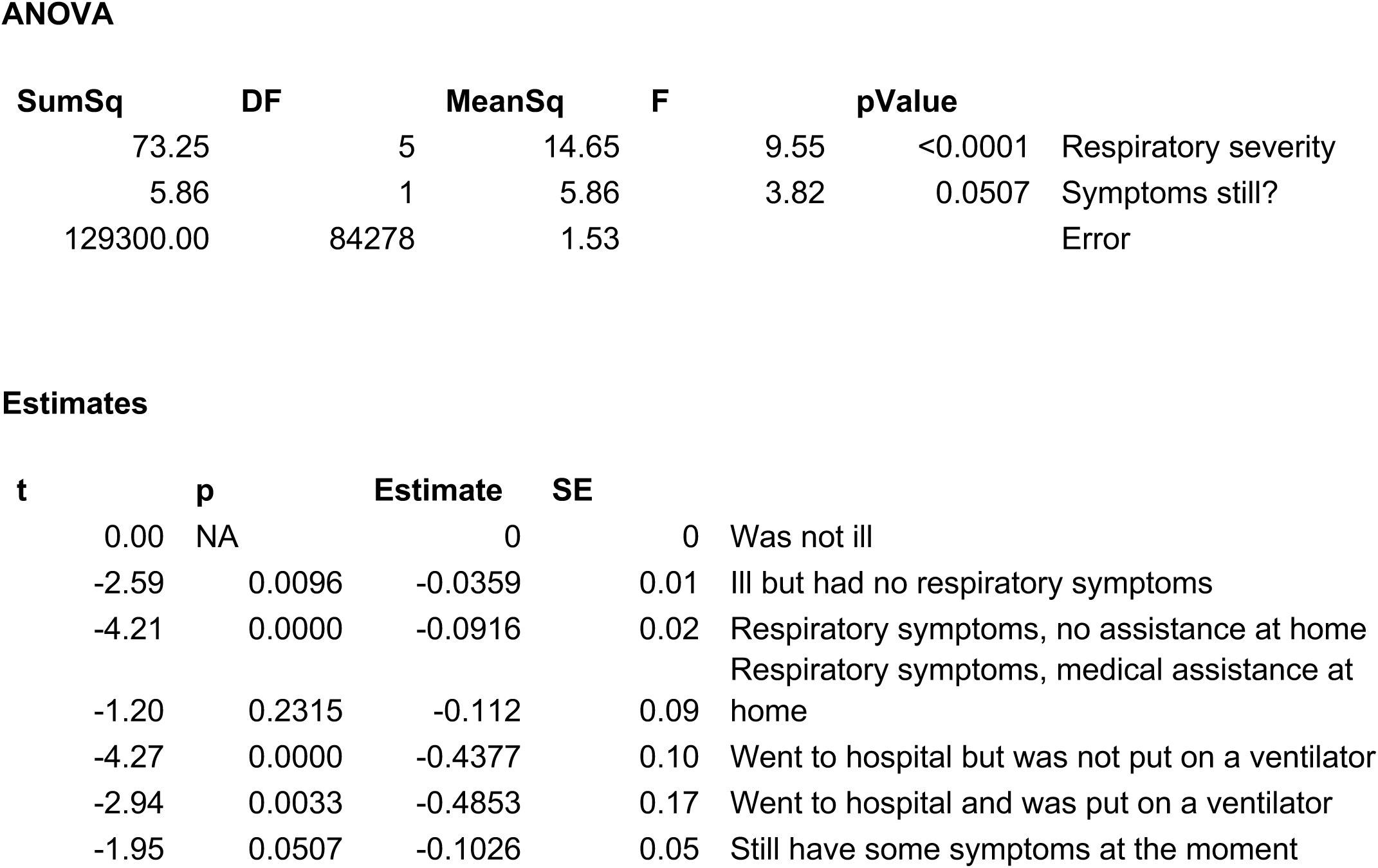
General linear model including residual symptoms as a factor

**Table S6.** GLM including pre-existing conditions

**Incidence**
|  |  |
| --- | --- |
| 2289 | **Weakened immune system** |
| 619 | **Kidney disease** |
| 3128 | **Diabetes** |
| 2385 | **Heart disease** |
| 822 | **High blood pressure** |
| 202 | **Heart problems** |
| 426 | **Liver disease** |
| 9115 | **Lung conditions** |
| 167 | **Spleen/sickle cell** |

**ANOVA**
| **SumSq** | **DF** | **MeanSq** | **F** | **pValue** | **Estimate** |  |
| --- | --- | --- | --- | --- | --- | --- |
| 17.18 | 1 | 17.18 | 11.20 | 0.0008 | -0.090 | **Weakened immune system** |
| 1.11 | 1 | 1.11 | 0.72 | 0.3946 | -0.044 | **Kidney disease** |
| 29.69 | 1 | 29.69 | 19.36 | 0.0000 | -0.104 | **Diabetes** |
| 16.74 | 1 | 16.74 | 10.91 | 0.0010 | -0.087 | **Heart disease** |
| 0.11 | 1 | 0.11 | 0.07 | 0.7927 | -0.014 | **High blood pressure** |
| 0.84 | 1 | 0.84 | 0.55 | 0.4595 | -0.067 | **Heart problems** |
| 8.56 | 1 | 8.56 | 5.59 | 0.0181 | -0.145 | **Liver disease** |
| 2.20 | 1 | 2.20 | 1.44 | 0.2310 | -0.017 | **Lung conditions** |
| 1.14 | 1 | 1.14 | 0.75 | 0.3880 | 0.080 | **Spleen/sickle cell** |
| 84.84 | 5 | 16.97 | 11.07 | 0.0000 |  | **Respiratory severity** |
| 129220.00 | 84270 | 1.53 |  |  |  | **Error** |

| **Estimate** | **SE** | **tStat** | **pValue** |  |
| --- | --- | --- | --- | --- |
| 0a | 0a | 0a | na | Was not ill |
| -0.04 | 0.01 | -3.07 | 0.002 | Ill but had no respiratory symptoms |
| -0.09 | 0.02 | -4.39 | 0.000 | Respiratory symptoms, no assistance at home |
| -0.11 | 0.09 | -1.22 | 0.224 | Respiratory symptoms, medical assistance at home |
| -0.44 | 0.10 | -4.26 | 0.000 | Went to hospital but was not put on a ventilator |
| -0.52 | 0.16 | -3.21 | 0.001 | Went to hospital and was put on a ventilator |

**Table S7a.** Test scores correlation matrix

| Rare word definition | Tower of London | Block rearrange | Digit span | Emotional face discrimination | Mental rotation | Spatial span | Target detection | Analogical reasoning |  |
| --- | --- | --- | --- | --- | --- | --- | --- | --- | --- |
|  | 0.17 | 0.14 | 0.19 | 0.14 | 0.09 | 0.09 | 0.08 | 0.37 | Rare word definition |
| 0.17 |  | 0.31 | 0.14 | 0.11 | 0.09 | 0.20 | 0.09 | 0.22 | Tower of London |
| 0.14 | 0.31 |  | 0.11 | 0.09 | 0.15 | 0.20 | 0.11 | 0.20 | Block rearrange |
| 0.19 | 0.14 | 0.11 |  | 0.08 | 0.10 | 0.18 | 0.08 | 0.26 | Digit span |
| 0.14 | 0.11 | 0.09 | 0.08 |  | 0.01 | 0.06 | 0.05 | 0.11 | Emotional face discrimination |
| 0.09 | 0.09 | 0.15 | 0.10 | 0.01 |  | 0.20 | 0.22 | 0.24 | Mental rotation |
| 0.09 | 0.20 | 0.20 | 0.18 | 0.06 | 0.20 |  | 0.12 | 0.19 | Spatial span |
| 0.08 | 0.09 | 0.11 | 0.08 | 0.05 | 0.22 | 0.12 |  | 0.13 | Target detection |
| 0.37 | 0.22 | 0.20 | 0.26 | 0.11 | 0.24 | 0.19 | 0.13 |  | Analogical reasoning |

Tests included in the battery were selected based on deliverability via Internet browsers and phones, sensitivity to population variables of interest such as age, and tendency to measure different as opposed to a single common aspect of cognitive ability.

**Table S7b.** Rotated PCA loadings for 9 tests

|  | Verbal<br>problem<br>solving | Attention | Spatial<br>working<br>memory |
| --- | --- | --- | --- |
| Analogical reasoning | **0.6** | **0.3** | 0.1 |
| Rare word definition | **0.6** | 0.1 | 0.1 |
| Digit span | **0.3** | 0.1 | 0.1 |
| Tower of London | 0.2 | 0.1 | **0.5** |
| Block rearrange | 0.1 | 0.2 | **0.5** |
| Spatial span | 0.1 | **0.3** | **0.3** |
| Target detection | 0.1 | **0.3** | 0.1 |
| Mental rotation | 0.1 | **0.7** | 0.0 |
| *Emotional face discrimination* | *0.2* | *0.0* | *0.1* |

Three simple and interpretable components are evident for the varimax rotated factor model. The Emotional Discrimination test measures an ability that has little overlap with the other 8 tests.

**Table S7c.**
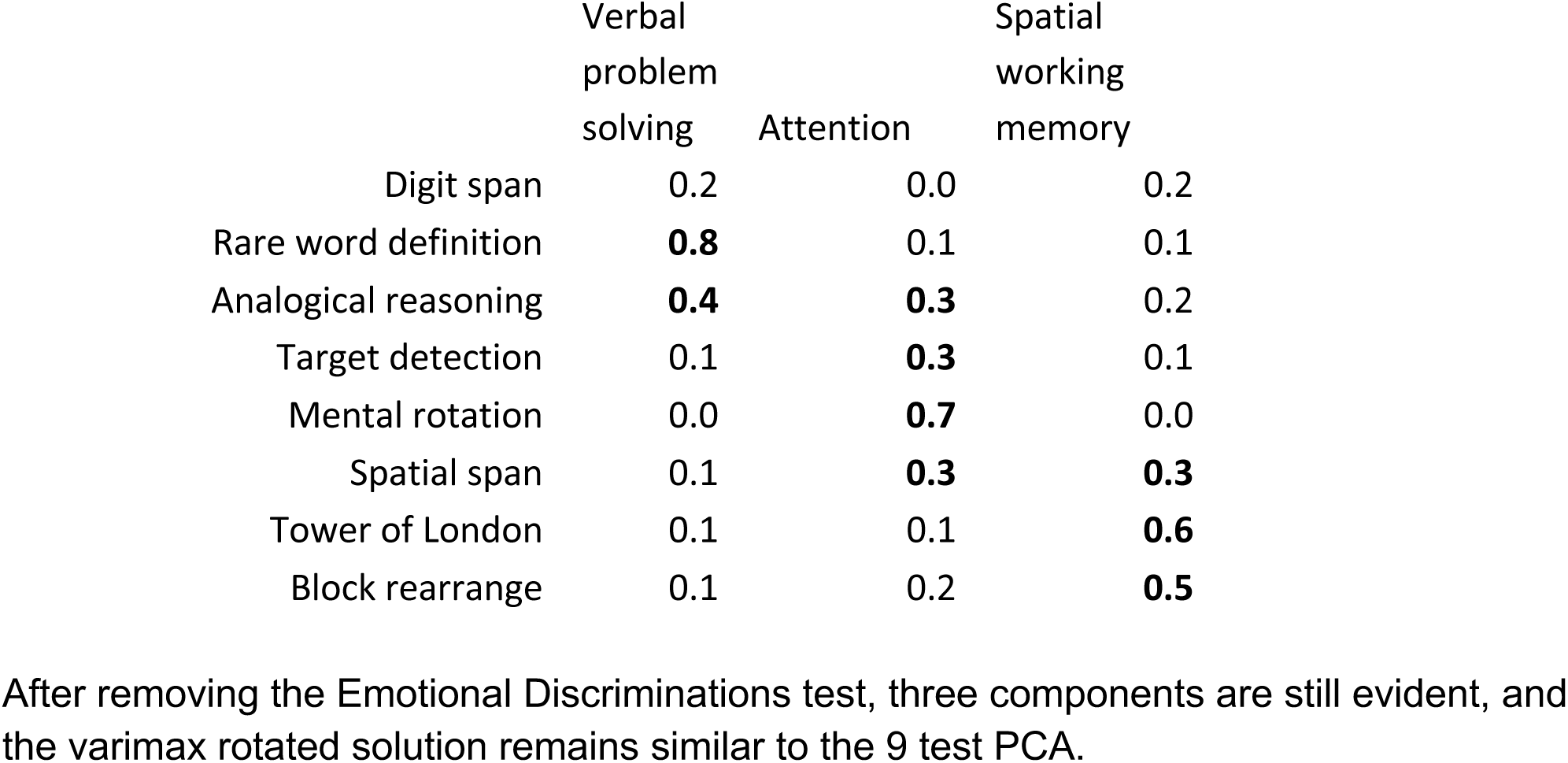
Rotated component loadings for 8 tests

|  | Verbal<br>problem<br>solving | Attention | Spatial<br>working<br>memory |
| --- | --- | --- | --- |
| Digit span | 0.2 | 0.0 | 0.2 |
| Rare word definition | **0.8** | 0.1 | 0.1 |
| Analogical reasoning | **0.4** | **0.3** | 0.2 |
| Target detection | 0.1 | **0.3** | 0.1 |
| Mental rotation | 0.0 | **0.7** | 0.0 |
| Spatial span | 0.1 | **0.3** | **0.3** |
| Tower of London | 0.1 | 0.1 | **0.6** |
| Block rearrange | 0.1 | 0.2 | **0.5** |

After removing the Emotional Discriminations test, three components are still evident, and the varimax rotated solution remains similar to the 9 test PCA.

**Table S7d.** GLMs of component scores vs respiratory severity measures

**ANOVAs**
|  | SumSq | DF | MeanSq | F | pValue |
| --- | --- | --- | --- | --- | --- |
| **COMPONENT 1** |  |  |  |  |  |
| Respiratory severity | 46 | 5 | 9.25 | 5.89 | <0.0001 |
| Error | 132350 | 84279 | 1.57 |  |  |
| **COMPONENT 2** |  |  |  |  |  |
| Respiratory severity | 71 | 5 | 14.29 | 7.46 | <0.0001 |
| Error | 161390 | 84279 | 1.92 |  |  |
| **COMPONENT 3** |  |  |  |  |  |
| Respiratory severity | 25 | 5 | 4.93 | 2.23 | 0.0489 |
| Error | 186830 | 84279 | 2.22 |  |  |

| Estimate | SE | tStat | pValue |  |
| --- | --- | --- | --- | --- |
| **COMPONENT 1** |  |  |  |  |
| 0.00 | 0.00 | 0.21 | 0.83 | (Intercept) |
| 0a | 0a | 0a | na | Was not ill |
| 0.02 | 0.01 | 1.30 | 0.19 | Ill but had no respiratory symptoms |
| -0.04 | 0.02 | -2.05 | 0.04 | Respiratory symptoms, no assistance at home |
| -0.09 | 0.09 | -0.95 | 0.34 | Respiratory symptoms, medical assistance at home |
| -0.28 | 0.10 | -2.74 | 0.01 | Went to hospital but was not put on a ventilator |
| -0.62 | 0.16 | -3.84 | 0.00 | Went to hospital and was put on a ventilator |
| **COMPONENT 2** |  |  |  |  |
| 0.01 | 0.01 | 2.05 | 0.04 | (Intercept) |
| 0a | 0a | 0a | na | Was not ill |
| -0.06 | 0.02 | -3.90 | 0.00 | Ill but had no respiratory symptoms |
| -0.07 | 0.02 | -2.85 | 0.00 | Respiratory symptoms, no assistance at home |
| -0.22 | 0.10 | -2.15 | 0.03 | Respiratory symptoms, medical assistance at home |
| -0.33 | 0.11 | -2.92 | 0.00 | Went to hospital but was not put on a ventilator |
| -0.33 | 0.18 | -1.86 | 0.06 | Went to hospital and was put on a ventilator |
| **COMPONENT 3** |  |  |  |  |
| 0.01 | 0.01 | 1.15 | 0.25 | (Intercept) |
| 0a | 0a | 0a | na | Was not ill |
| -0.04 | 0.02 | -2.51 | 0.01 | Ill but had no respiratory symptoms |
| -0.04 | 0.03 | -1.54 | 0.12 | Respiratory symptoms, no assistance at home |
| 0.07 | 0.11 | 0.62 | 0.54 | Respiratory symptoms, medical assistance at home |
| -0.15 | 0.12 | -1.22 | 0.22 | Went to hospital but was not put on a ventilator |
| -0.21 | 0.19 | -1.10 | 0.27 | Went to hospital and was put on a ventilator |

**Table S8.** Analysis of individual test summary scores relative to respiratory severity

|  | Rare word definitions | Analogical reasoning | Target Detection | Tower of London | Mental rotation | Block rearrange | Spatial span | Emotional discrimination | Digit span |
| --- | --- | --- | --- | --- | --- | --- | --- | --- | --- |
| **ANOVA P** | <0.0001 | <0.0001 | 0.0003 | 0.0089 | <0.0001 | 0.0082 | 0.0006 | 0.0006 | 0.0297 |
| **Task coefficients relative to control** |  |  |  |  |  |  |  |  |  |
| Not ill | 0.00 | 0.00 | 0.00 | 0.00 | 0.00 | 0.00 | 0.00 | 0.00 | 0.00 |
| No respiratory symptoms | 0.00 | -0.02 | -0.02 | -0.03 | -0.04 | -0.04 | -0.04 | 0.04 | 0.01 |
| No assistance at home | -0.04 | -0.06 | -0.04 | -0.03 | -0.04 | -0.02 | -0.06 | -0.02 | -0.04 |
| Assistance at home | -0.09 | -0.03 | -0.13 | -0.01 | -0.15 | 0.04 | -0.10 | -0.07 | -0.02 |
| Hospital, no ventilator | -0.22 | -0.38 | -0.16 | -0.11 | -0.23 | -0.15 | -0.06 | -0.18 | 0.00 |
| Hospital with ventilator | -0.53 | -0.42 | -0.35 | -0.29 | -0.24 | -0.17 | -0.10 | -0.01 | 0.32 |
| **Task p relative to control** |  |  |  |  |  |  |  |  |  |
| Not ill | 1 | 1 | 1 | 1 | 1 | 1 | 1 | 1 | 1 |
| No respiratory symptoms | 0.7502 | 0.0804 | 0.0423 | 0.0175 | 0.0003 | 0.0028 | 0.0018 | 0.0003 | 0.3511 |
| No assistance at home | 0.0178 | 0.0009 | 0.0158 | 0.0684 | 0.0117 | 0.2917 | 0.0011 | 0.2006 | 0.0325 |
| Assistance at home | 0.2260 | 0.6961 | 0.0812 | 0.9244 | 0.0420 | 0.5875 | 0.1919 | 0.3868 | 0.8456 |
| Hospital, no ventilator | 0.0069 | <0.0001 | 0.0541 | 0.1252 | 0.0043 | 0.0567 | 0.4369 | 0.0275 | 0.9224 |
| Hospital with ventilator | <0.0001 | 0.0011 | 0.0056 | 0.0262 | 0.0596 | 0.1361 | 0.4245 | 0.9025 | 0.0103 |

